# The effects of inhaled corticosteroids on healthy airways

**DOI:** 10.1101/2023.11.14.23298442

**Authors:** Emanuele Marchi, Timothy S.C. Hinks, Matthew Richardson, Latifa Khalfaou, Fiona A. Symon, Poojitha Rajasekar, Rachel Clifford, Beverley Hargadon, Cary D. Austin, Julia L. MacIsaac, Michael S. Kobor, Salman Siddiqui, Jordan S. Mar, Joseph R. Arron, David Choy, Peter Bradding

## Abstract

**Rationale:** The effects of inhaled corticosteroids (ICS) on healthy airways are poorly defined.

**Objectives:** To delineate the effects of ICS on gene expression in healthy airways, without confounding caused by changes in disease-related genes and disease-related alterations in ICS-responsiveness.

**Methods:** Randomised open-label bronchoscopy study of high dose ICS therapy in 30 healthy adult volunteers randomised 2:1 to i) fluticasone propionate 500 mcg bd or ii) no treatment, for 4 weeks. Laboratory staff were blinded to allocation. Biopsies and brushings were analysed by immunohistochemistry, bulk RNA sequencing, DNA methylation array and metagenomics.

**Measurements and main results:** ICS induced small between-group differences in blood and lamina propria eosinophil numbers, but not in other immunopathological features, blood neutrophils, FeNO, FEV_1_, microbiome or DNA methylation. ICS treatment upregulated 72 genes in brushings and 53 genes in biopsies, and downregulated 82 genes in brushings and 416 genes in biopsies. The most downregulated genes in both tissues were canonical markers of type-2 inflammation (FCER1A, CPA3, IL33, CLEC10A, SERPINB10 and CCR5), T cell-mediated adaptive immunity (TARP, TRBC1, TRBC2, PTPN22, TRAC, CD2, CD8A, HLA-DQB2, CD96, PTPN7), B cell immunity (CD20, immunoglobulin heavy and light chains), and innate immunity, including CD48, Hobit, RANTES, Langerin and GFI1. An IL-17-dependent gene signature was not upregulated by ICS.

**Conclusions:** In healthy airways, 4-week ICS exposure reduces gene expression related to both innate and adaptive immunity, and reduces markers of type-2 inflammation. This implies that homeostasis in health involves tonic type-2 signalling in the airway mucosa, which is exquisitely sensitive to ICS.

## INTRODUCTION

Inhaled corticosteroids (ICS) are the cornerstone of asthma treatment. They attenuate eosinophilic airway inflammation(1,2), improve lung function, and reduce asthma symptoms, exacerbations, and mortality(3). However, their use is also associated with an increased risk of pneumonia(4). Corticosteroids modulate the expression of many molecular pathways at the level of gene transcription, through direct upregulation of anti-inflammatory molecules and β-adrenoceptors (transactivation), and suppression of pro-inflammatory genes, either through direct DNA-binding or via inhibition of pro-inflammatory transcription factor binding (transrepression)(5). In people with severe asthma, ICS are relatively ineffective even at high doses, but the mechanisms behind this corticosteroid insensitivity are poorly understood and likely multi-factorial(6,7).

When considering the underlying molecular pathways driving both severe asthma and relative corticosteroid insensitivity, it is unclear to what extent this is driven by pathways that are not responsive to corticosteroids, as opposed to inhibition of corticosteroid signalling. Gene expression profiling in asthmatic epithelial bronchial brushings and bronchial biopsies has identified several molecular pathways present in subgroups of patients with mild, moderate and severe asthma(8,9). Approximately 80% of people with steroid-naïve “mild” asthma demonstrate evidence of blood or airway eosinophilia, with a concomitant increase in the fraction of exhaled nitric oxide (FeNO)(10,11). This phenotype is characterised by increased expression of an airway gene expression signature driven by IL-4 and IL-13(8,9,12,13); tissue eosinophilia is also dependent on IL-5(14). Together these cytokines are described as Th2 or type-2 cytokines (T2). T2 expression and the accompanying eosinophilia are suppressed by ICS in mild asthma(15). In severe asthma, a persistent T2 gene signature is evident in about 25% of patients, suggesting corticosteroid insensitivity(8,9). In addition, in severe asthma, about 25% of patients have evidence of an IL-17-dependent gene signature, which is seen only in people on ICS(8,9), and mutually exclusive with the T2 signature. It is therefore not clear whether this IL-17 activity represents an independent corticosteroid-insensitive pathway driving severe asthma or a consequence of ICS therapy. Approximately 50% of people with severe asthma have neither a T2- or IL-17-dependent airway gene signature, and the mechanisms driving their persistent disordered airway physiology remains unknown. So, while gene expression profiling provides insight into the abnormalities present in severe asthmatic airways, the multiple effects of ICS on airway gene expression make it difficult to disentangle the changes due to the disease as opposed to the treatment, and whether corticosteroid activity is inhibited or not.

The effects of ICS on healthy airways are poorly defined. We hypothesise that gene expression data in severe asthma will be more interpretable if we can delineate and thus allow for the effects of high dose ICS therapy. We have therefore performed a randomised open label bronchoscopy study of high dose ICS therapy in healthy adult volunteers, with the aim of understanding transcriptional consequences of ICS therapy without the confounding effects due to disease-related processes.

## METHODS

Detailed methods are provided in the online data supplement.

This prospective study was approved by the East Midlands-Leicester Central Research Ethics Committee (reference:15/EM/0313) and registered at clinicaltrials.gov (NCT02476825). Participants gave written informed consent.

### Participant population

Healthy volunteers aged 18-65 were eligible, were current non-smokers with <10 pack year smoking history, and had no prior history or clinical evidence of lower respiratory disease with normal spirometry. Participants with a history of rhinitis were required to have a PC_20_ methacholine >16 mg/ml.

### Study design

This was a randomised, open-labelled, bronchoscopy study designed to assess the effects of 4 weeks treatment with fluticasone propionate on airway gene expression and cellularity in healthy adult volunteers. The primary endpoint was the corticosteroid-inducible gene expression pattern in healthy airways. Secondary endpoints included the relative change from baseline in airway cellularity.

30 participants were randomised by a blinded investigator (MR) in a 2:1 ratio to one of two study groups: i) fluticasone propionate 500 mcg b.i.d. via Accuhaler (Diskus) for 4 weeks (n=20), or ii) no treatment (observation) for 4 weeks (n=10). Bronchoscopy was performed at baseline prior to the start of treatment/observation, and at the end of week 4. Genentech and Leicester laboratory support staff were blinded to treatment allocation.

To ensure there were sufficient data for analysis, if a subject withdrew before completion of the study, a further subject(s) was randomised after the first 30 randomisations until a total of 30 subjects had completed the study.

### Bronchoscopy

Subjects underwent bronchoscopy conducted according to British Thoracic Society guidelines(16). Mucosal biopsies and brushes were collected from 2^nd^-5^th^ generation bronchi under direct vision as per study procedure manual.

### Tissue processing, immunohistochemistry and assessment of immunopathology

Please see the online supplement.

All pathological data were assessed by an observer blinded to the identity and treatment allocations of the participants.

### RNA sequencing

Please see the online supplement.

### Bisulphite conversion and DNA methylation arrays, DNA methylation data quality control and normalization, Differential DNA methylation analysis, Expression quantitative trait methylation (eQTM) analysis

Please see the online supplement.

### Microbiota Sequence Data Generation, Processing, and Analysis

Please see the online supplement.

### Transcriptomic analysis

Sequences in fastq files (in single and pair ends) were aligned using STAR aligner (version 2.7.1a) to the human reference genome GRCh38; R package (17)Rsubread was employed for quantification of reads assigned to genes.

Raw count pre-processing, normalisation and differential gene expression analysis was performed using R packaged DESeq2, edgeR, and limma. Gene pathways enrichment analysis was executed using R packages fgsea, clusterProfile, and ReactomePA. Heatmaps, volcano and MA plots were generated using in-home R code, ggplot2 and plotly packages(17,18).

### Statistical analysis

Basic summary statistical analysis was performed using GraphPad Prism version 7.03 (GraphPad Software, San Diego). Parametric and non-parametric data are presented as mean (standard deviation [SD]) and median (interquartile range [IQR]) respectively unless otherwise stated.

## RESULTS

### Clinical characteristics

We recruited 44 healthy participants. 32 proceeded to bronchoscopy but one patient withdrew from the study after the first bronchoscopy. 31 completed 2 bronchoscopies, but one was subsequently withdrawn due to both <80% medication adherence and an intercurrent asymptomatic bronchitis evident at the 2^nd^ bronchoscopy (with bronchial wash samples positive for a non-Covid coronavirus and Staphylococcus aureus). The clinical characteristics of the 30 participants completing the study are shown in **table 1**.

**Table 1.**
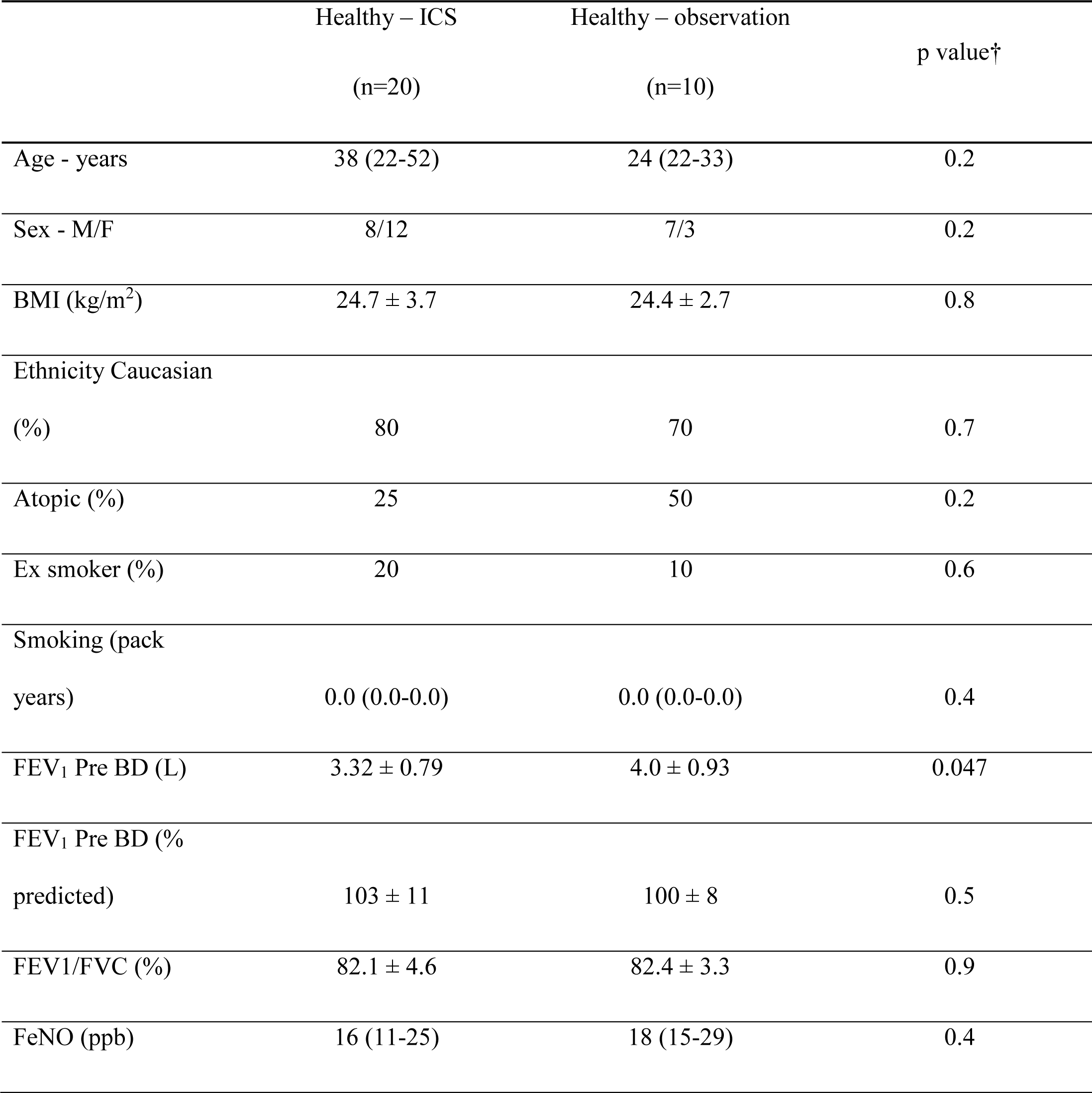
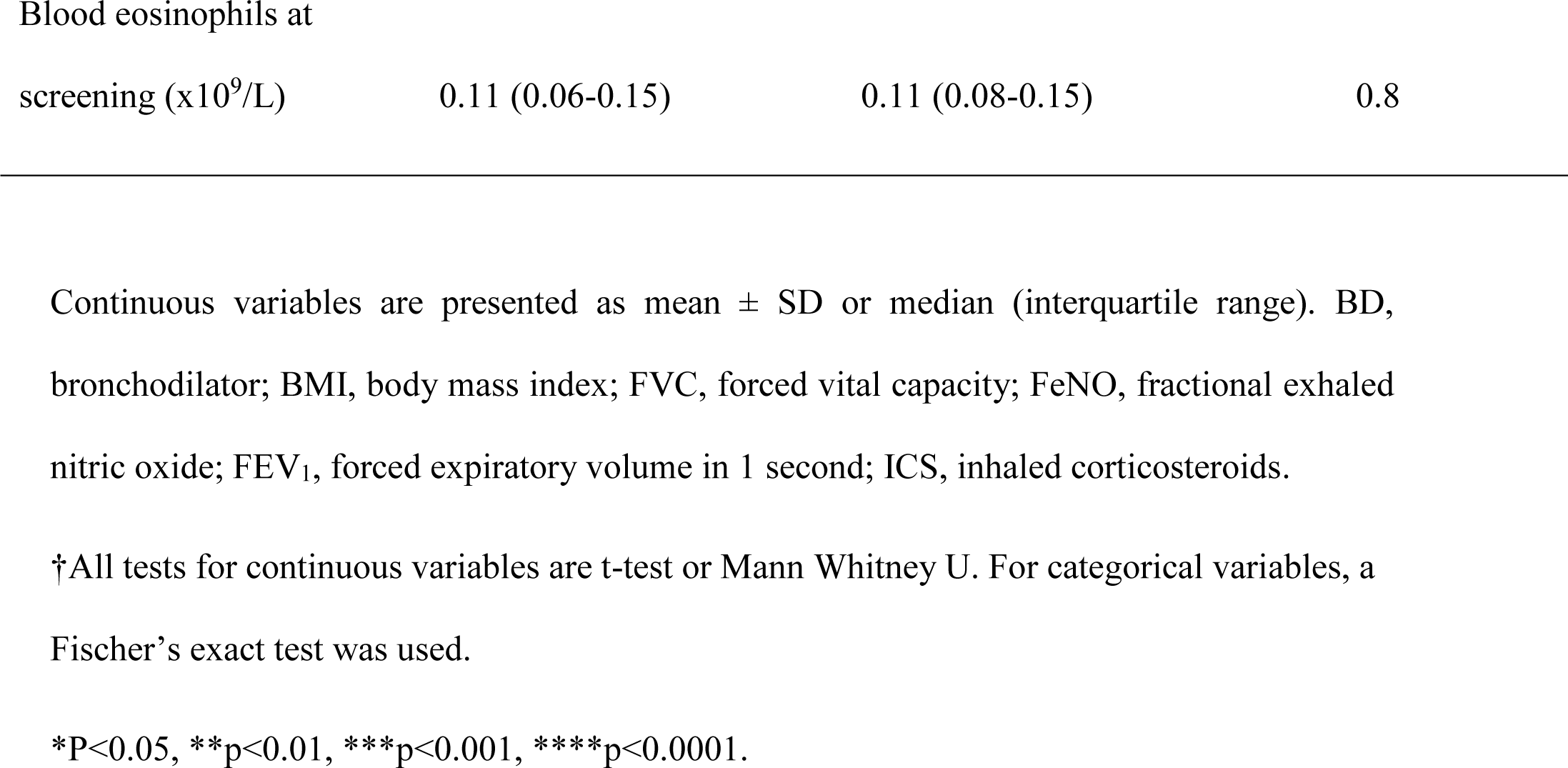
Demographics of the study participants.

### Effects of ICS on biomarker and physiological measurements

There was a significant increase in blood eosinophil numbers after 4 weeks in the observation group compared to people using ICS, but this was not related to atopic status (**Supplementary figure E1A**). There were no significant between-group differences for changes in blood neutrophils, FeNO or FEV_1_ (**Supplementary figure E1B-D**).

### Effects of ICS on airway inflammatory and structural cells

Suitable paired samples for immunohistochemical analysis of the lamina propria were available from 17 participants receiving ICS and 8 undergoing observation (**Figure 1** and **Supplementary Figure E2**). Although there was no significant change in lamina propria eosinophil counts within either group from the 1^st^ to 2^nd^ bronchoscopy, there was a significant between-group difference in the changes (p=0.01), due to a non-significant increase in the observation group (**Figure 1A**). There was no correlation between the change in blood eosinophils versus tissue eosinophils in the observation group (r_s_= −0.024, p=0.97). Comparing the changes in measurements from the 1^st^ to 2^nd^ bronchoscopy for the ICS versus the observation group, there were no significant differences between treatment groups for lamina propria neutrophil, tryptase+ or chymase+ mast cell counts, airway smooth muscle (ASM) and epithelial area expressed as a percentage of biopsy area, or reticular basement membrane depth (**Figure 1B-G)**(**Supplementary Figure E2**).

**Figure 1.**
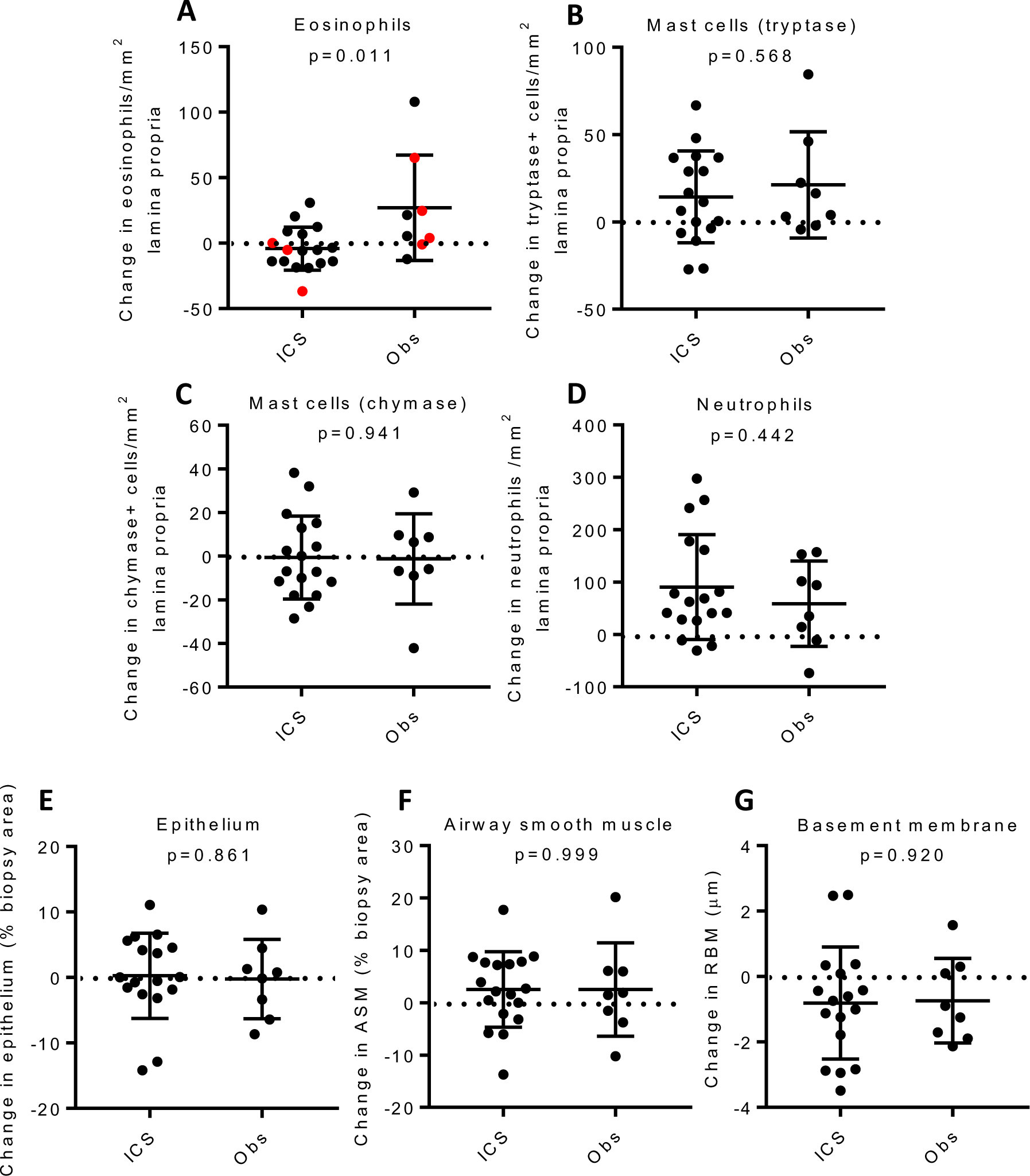
Immunohistochemical analysis of the lamina propria biopsies showing cell counts and remodelling features on participants with available paired data, before and after 4 weeks treatment with inhaled fluticasone or without treatment. The change in numbers of **A**) eosinophils, with atopic participants shown in red, **B**) tryptase-positive mast cells, **C**) chymase-positive mast cells, or **D**) neutrophils, expressed in absolute counts/mm^2^. Changes in area of **E**) epithelium or **F**) airway smooth muscle (ASM), expressed as a percentage of biopsy area or of **G**) reticular basement membrane (RBM) thickness. Horizontal bars represent mean (SD)(mast cells, neutrophils, ASM, epithelium, RBM) or median (IQR) (eosinophils), analysed by unpaired t test or Mann Whitney U respectively.

### Effects of ICS on airway gene expression

Suitable paired brush and biopsy samples were available for 15 and 20 participants respectively receiving ICS. We observed significant differential expression of genes amongst participants at week 4 compared with baseline in the ICS-treatment group, with upregulation of 72 genes in brushings and 53 genes in biopsies, and downregulation of 82 genes in brushings and 416 genes in biopsies (**Figure 2**, **Supplementary Tables E1-E4, Supplementary Figures E3, E4**). Amongst participants in the observation-only group there were no significant changes in gene expression observed between baseline and week 4 (**Supplementary Figure E5**). There was a close correlation between epithelial brush and bronchial biopsy gene expression, with 20 genes common to the top 24 most significantly differentially upregulated genes in both airway compartments (**Table 2**) and 20 genes common to the top 41 most significantly downregulated genes in both compartments (**Table 3**).

**Figure 2.**
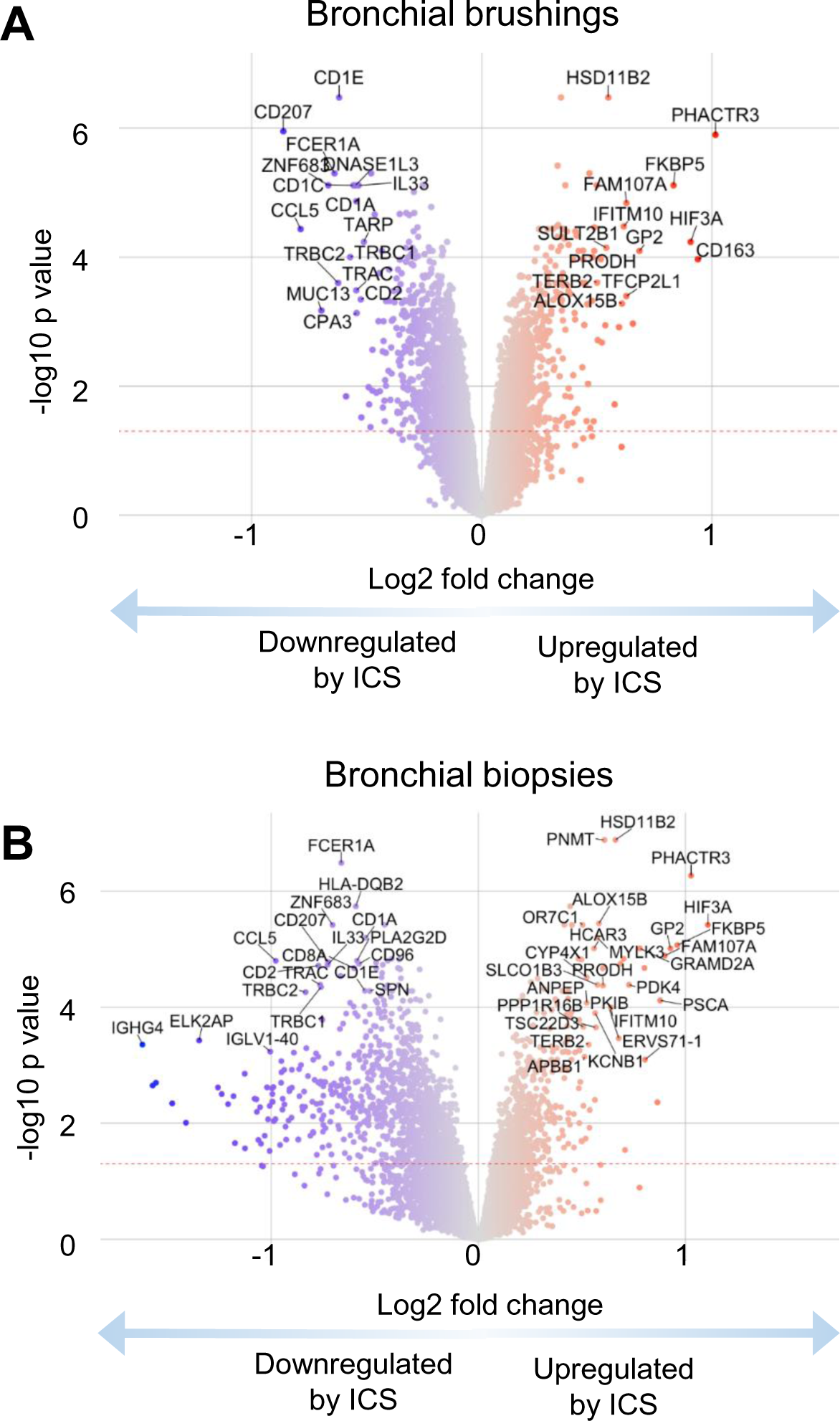
Changes in gene expression measured by RNAseq in response to 4 weeks treatment with inhaled fluticasone. **A**) Bronchial brush volcano plot. **B**) Bronchial biopsy volcano plot.

**Table 2.** Top upregulated differentially expressed genes.

| Gene | Bronchial brushes |  |  | Bronchial biopsies |  |  | Mean of FDR<br>P values | Comments and aliases | Functional role |
| --- | --- | --- | --- | --- | --- | --- | --- | --- | --- |
|  | Baseline<br>read count | Log2 fold<br>change | FDR P value | Baseline<br>read count | Log2 fold<br>change | FDR P<br>value |  |  |  |
| PHACTR3 | 497 | 3.59 | 1.43E-41 | 325 | 3.28 | 4.29E-38 | 2.14E-38 | Phosphatase And Actin Regulator 3; associated with the nuclear scaffold in proliferating cells. | Cytoskeletal changes |
| HSD11B2 | 468 | 1.89 | 3.22E-41 | 392 | 1.81 | 9.88E-37 | 4.94E-37 | Hydroxysteroid 11-β Dehydrogenase 2: converts cortisol to cortisone, prevents mineralocorticoid receptor activation | Steroid metabolism |
| FKBP5 | 3336 | 1.88 | 2.17E-27 | 5029 | 1.67 | 3.14E-20 | 1.57E-20 | Regulates corticosteroid sensitivity; asthma susceptibility gene. | Steroid metabolism |
| FAM107A | 3486 | 1.45 | 6.14E-22 | 3657 | 1.60 | 4.02E-17 | 2.01E-17 | Family With Sequence Similarity 107 Member A; role in actin and microtubule organisation. | Cytoskeletal changes |
| HIF3A | 584 | 3.18 | 4.42E-17 | 990 | 2.67 | 5.35E-21 | 2.21E-17 | Hypoxia Inducible Factor 3 Subunit $\alpha$; canonical sensor of hypoxia. | Hypoxic sensing |
| MYLK3 | 476 | 1.10 | 1.19E-40 | 490 | 1.47 | 6.54E-17 | 3.27E-17 | Myosin Light Chain Kinase 3; Immune response to CCR3 signaling in eosinophils, regulates smooth muscle contracti | Smooth muscle/immune |
| GP2 | 341 | 2.48 | 1.26E-15 | 286 | 3.12 | 9.81E-28 | 6.29E-16 | Glycoprotein 2; binds pathogens such as enterobacteria. | Innate immune |
| SULT2B1 | 1706 | 1.38 | 3.05E-17 | 1199 | 1.44 | 2.01E-15 | 1.02E-15 | Hydroxysteroid Sulfotransferase 2; catalyses the sulfate conjugation hormones. | Steroid metabolism |
| GRAMD2A | 2452 | 1.15 | 5.37E-24 | 2309 | 1.38 | 2.69E-15 | 1.35E-15 | GRAM Domain Containing 2A; organization of endoplasmic reticulum-plasma membrane contact sites. | Cellular metabolism |
| SYT8 | 503 | 1.12 | 2.93E-15 | 470 | 1.13 | 2.37E-22 | 1.46E-15 | Synaptotagmin 8; regulate exocytosis of hormones and in intracellular organelles. | Steroid metabolism |
| SLCO1B3 | 124 | 1.94 | 1.69E-17 | 111 | 2.27 | 5.11E-14 | 2.55E-14 | Solute Carrier Organic Anion Transporter 1B3; transports conjugated steroids, leukotrienes, prostaglandins. | Steroid metabolism |
| HCAR2 | 4673 | 1.10 | 9.06E-20 | 3361 | 1.22 | 1.15E-13 | 5.73E-14 | Niacin Receptor 1; receptor for niacin and mediates neutrophil apoptosis and anti-inflammatory effect of butyrate | Anti-inflammatory |
| PSCA | 23905 | 1.21 | 9.32E-14 | 13008 | 1.87 | 3.83E-14 | 6.58E-14 | Prostate stem cell antigen; may regulate cell proliferation. | Cell proliferation |
| TFCP2L1 | 3228 | 1.38 | 2.76E-13 | 4849 | 1.40 | 1.64E-16 | 1.38E-13 | Transcription corepressor activity, and probably Wnt/β-catenin signaling. | Cell proliferation |
| PRODH | 5268 | 1.20 | 5.53E-14 | 4754 | 1.10 | 8.22E-13 | 4.39E-13 | Proline Dehydrogenase 1; catalyses proline catabolism. | Cellular metabolism |
| TSC22D3 | 7099 | 1.12 | 1.19E-17 | 8967 | 1.14 | 1.55E-12 | 7.73E-13 | Glucocorticoid-Induced Leucine Zipper; mediates anti-inflammatory effects of glucocorticoids in macrophages. | Anti-inflammatory |
| IFITM10 | 1872 | 1.55 | 4.46E-22 | 1816 | 1.23 | 3.71E-11 | 1.86E-11 | IFITM family functions include controlling cell proliferation, and promoting homotypic cell adhesion. | Cell proliferation |
| ANPEP | 2159 | 1.16 | 4.30E-14 | 1707 | 1.02 | 6.28E-11 | 3.14E-11 | Aminopeptidase M, CD13; broad specificity aminopeptidase, promotes angiogenesis, and cell growth. | Cell proliferation |
| KCNB1 | 602 | 1.03 | 1.83E-15 | 662 | 1.28 | 6.63E-10 | 3.31E-10 | Potassium Voltage-Gated Channel Subfamily B Member 1; neurotransmission and regulation of exocytosis. | Cell signalling |
| GNMT | 704 | 1.28 | 2.24099E-12 | 548 | 1.10 | 1.97E-07 | 9.84E-08 | Glycine N-Methyltransferase; regulation of methyl group metabolism. | Cellular metabolism |

Top 20 upregulated genes common to both epithelial brushes and bronchial biopsies occurring in the top 24 differentially upregulated genes
when ranked by FDR p value. Table ranks genes by arithmetic mean of FDR p values across both tissues.

**Table 3.** Top downregulated differentially expressed genes.

| Gene | Bronchial brushes |  |  | Bronchial biopsies |  |  | Mean of FDR<br>P values | Comments and aliases | Functional role |
| --- | --- | --- | --- | --- | --- | --- | --- | --- | --- |
|  | Baseline<br>read count | Log2 fold<br>change | FDR P<br>value | Baseline<br>read count | Log2 fold<br>change | FDR P<br>value |  |  |  |
| FCER1A | 144 | -2.78 | 2.15E-24 | 228 | -2.25 | 6.93E-29 | 1.08E-24 | High affinity IgE receptor; expressed on airway mast cells and dendritic cells. | Type 2 immunity |
| ZNF683 | 136 | -3.33 | 2.31E-23 | 131 | -3.06 | 1.32E-20 | 6.62E-21 | Hobit; transcription factor promoting tissue residency in innate and adaptive lymphocytes. | Adaptive immunity |
| CD207 | 169 | -3.78 | 3.05E-23 | 144 | -2.90 | 4.24E-18 | 2.12E-18 | Langerin; C-type lectin with mannose binding specificity, involved in nonclassical antigen-processing. | Antigen presentation |
| CCL5 | 1133 | -2.39 | 1.21E-15 | 1233 | -2.15 | 2.67E-16 | 7.37E-16 | RANTES; chemoattractant for monocytes, memory T helper cells and eosinophils. | Adaptive immunity |
| IL33 | 3885 | -1.20 | 2.18E-22 | 6985 | -1.24 | 3.70E-15 | 1.85E-15 | Interleukin 33; airway epithelial cell alarmin which acts via ST2 to promote type-2 responses. | Alarmin |
| CD96 | 253 | -1.62 | 1.31E-12 | 399 | -1.64 | 3.49E-14 | 6.71E-13 | TACTILE (T cell activation, increased late expression); involved in adhesion of T and NK cells to target cells. | Cell mediated immunity |
| TARP | 100 | -2.50 | 7.30E-15 | 112 | -1.90 | 3.18E-10 | 1.59E-10 | TCR Gamma Alternate Reading Frame Protein. | T cell adaptive immunity |
| TRBC1 | 353 | -2.33 | 3.39E-10 | 540 | -2.07 | 3.65E-13 | 1.70E-10 | T cell receptor Beta constant chain 1. | T cell adaptive immunity |
| TRBC2 | 463 | -2.41 | 7.16E-10 | 741 | -2.07 | 2.98E-13 | 3.58E-10 | T cell receptor Beta constant chain 2. | T cell adaptive immunity |
| PTPN22 | 165 | -1.26 | 1.43E-09 | 243 | -1.52 | 5.76E-11 | 7.42E-10 | Protein Tyrosine Phosphatase Non-Receptor Type 22. Regulates Treg frequency and T cell motility. | T cell adaptive immunity |
| TRAC | 539 | -1.92 | 3.37E-09 | 764 | -1.84 | 4.48E-13 | 1.68E-09 | T cell receptor Alpha constant chain. | T cell adaptive immunity |
| CPA3 | 228 | -2.26 | 7.00E-09 | 913 | -1.98 | 1.51E-09 | 4.26E-09 | Carboxypeptidase A3; mast cell-specific peptidase located in secretory granules. | Type 2 immunity |
| CD2 | 566 | -1.87 | 1.23E-08 | 732 | -1.87 | 9.60E-14 | 6.14E-09 | Lymphocyte-Function Antigen-2; T and NK cell marker and co-stimulatory molecule. | T cell adaptive immunity |
| CLEC10A | 135 | -1.71 | 1.82E-08 | 199 | -2.20 | 4.39E-14 | 9.11E-09 | C-Type Lectin Domain Containing 10A; CD301: marker for alternatively activated macrophages. | Type 2 immunity |
| SERPINB10 | 287 | -1.18 | 1.82E-08 | 350 | -1.42 | 1.25E-12 | 9.12E-09 | Serpin Family B Member 10; inhibits Th2 cell apoptosis, highly upregulated on airway epithelium by IL-13. | Type 2 immunity |
| CD8A | 516 | -1.55 | 6.40E-07 | 611 | -1.52 | 5.11E-14 | 3.20E-07 | T cell co-receptor for MHC1 restriction. | T cell adaptive immunity |
| GFI1 | 133 | -1.06 | 1.73E-06 | 177 | -1.23 | 2.46E-10 | 8.64E-07 | Growth Factor Independent 1 Transcriptional Repressor; involved in haematopoiesis, especially of neutrophils. | Innate immunity |
| HLA-DQB2 | 798 | -1.01 | 9.52E-06 | 640 | -1.51 | 4.63E-18 | 4.76E-06 | Major histocompatibility complex, class II, DQ beta 2 | T cell adaptive immunity |
| CCR5 | 295 | -1.42 | 9.97E-05 | 426 | -1.91 | 1.11E-12 | 4.99E-05 | Ligands include CCL5 (RANTES), MCP-2, MIP-1a, MIP-1b. Chemotactic for T2-induced eosinophils. | Type 2 immunity |
| PTPN7 | 248 | -1.32 | 0.00033 | 312 | -1.64 | 2.36E-10 | 1.67E-04 | Protein Tyrosine Phosphatase Non-Receptor 7; regulation of TCR signaling, early response gene in lymphokine stimulated T cell adaptive immunity | T cell adaptive immunity |

Top 20 downregulated genes common to both epithelial brushes and bronchial biopsies occurring in the top 41 biopsy differentially
downregulated genes when ranked by FDR p value. Table ranks genes by arithmetic mean of FDR p values across both tissues.

The most significantly upregulated genes were predominantly those involved in steroid metabolism (HSD11B2, FKBP5, SULT2B1, SYT8, and SLCO1B3), cellular proliferation (PSCA, TFCP2L1, IFITM10, ANPEP), cellular metabolism (GRAMD2A, PRODH, GNMT) and cytoskeletal changes (PHACTR3, FAM107A). By contrast genes which were most significantly downregulated in both brushings and biopsies were key components of T2-driven inflammation (FCER1A, CPA3, IL33, CLEC10A, SERPINB10 and CCR5) and T cell-mediated adaptive immunity (TARP, TRBC1, TRBC2, PTPN22, TRAC, CD2, CD8A, HLA-DQB2, CD96, PTPN7) and (**Table 3**). All other top 20 common downregulated genes were involved with innate or adaptive immunity, including the transcription factor Hobit (ZNF683) which promotes lymphocyte tissue residency, the chemokine RANTES (CCL5), the antigen presentation-associated molecule Langerin (CD207), and the growth factor GFI1 which is involved in haematopoiesis, especially of neutrophils. In addition, there was downregulation of genes associated with B cell function and immumunoglobulin production (CD20/79, most heavy and variable light chains for IgA, IgG and IgM, JCHAIN), protective innate immunity (e.g. CD48, CD163), mast cell proteases (TPSB1, TPSAB1, CPA3), the beta chain of the high affinity IgE receptor (MS4A2), and prostaglandin D2 synthase (PTGDS1).

Consistent with these findings, pathway analysis of bronchial brushings with Reactome showed strong ICS-related downregulation of innate and adaptive pathways, including ‘*Immunoregulatory interactions between a lymphoid and non-lymphoid cell*’, five pathways related to TCR signaling, the immunological synapse or co-stimulation, with weaker signals for ‘*Generation of second messenger molecules’, ‘PD-1 signaling’,* and *‘Chemokine receptors bind chemokines*’, showing the potent ability of ICS to suppress local adaptive T cell immunity (**Supplementary Figures E6, Supplementary Table E5**). Parallel analysis of the bronchial biopsies again showed effects on lymphoid – non-lymphoid interactions and TCR signaling, but also strong suppression of ‘*Extracellular matrix organization’, ‘Cell surface interactions at the vascular wall’* and *‘Integrin cell surface interactions’,* suggesting the potential for suppression of inflammatory cell recruitment to the mucosa (**Supplementary Figure E7, Supplementary Table E6**). ICS did not upregulate the IL-17-dependent gene signature identified previously in people with moderate-severe asthma (**Supplementary Fig.E8**)

Next we compared our set of differentially upregulated genes with a set of 26 genes previously reported as induced by 10 weeks of inhaled fluticasone in bronchial brushings from participants with mild asthma(19). Geneset enrichment analysis profiles showed very close agreement between genesets (**Supplementary Figure E9**), particularly for brushings, which were directly comparable between studies, showing very similar ICS-induced gene induction in health and mild asthma.

### Effects of ICS on the airway microbiome

Minimal effects of ICS treatment were observed on the airway microbiome (details in Supplementary Results and **Supplementary Figure E10**).

### Effects of ICS on airway DNA methylation

There were minimal effects of ICS treatment on DNA methylation (details in Supplementary Results and **Supplementary Figure E11**).

## DISCUSSION

In this open label, randomised study in healthy adult volunteers, we have defined genes that are altered directly by medium-term (4 weeks), twice daily high-dose ICS therapy independent of the confounding effect of asthma pathophysiology. Changes in airway cellularity were minimal between ICS treatment and no treatment, and there were no measurable biologically significant effects on the airway microbiome or DNA methylation.

By contrast, we observed widespread changes in gene expression following ICS use in these healthy individuals, which predominantly comprised downregulated genes. A striking finding was that the most significantly downregulated genes statistically were canonical markers of T2-driven inflammation. Notably the most highly downregulated gene, in both airway brushes, which predominantly sample the airway epithelium, and mucosal biopsies, was FCER1A which encodes the α chain of the high affinity IgE receptor, a key effector component of mast cell activation by allergens and of T2 immunity, which is highly upregulated in asthma (20,21). In addition, there was downregulation of genes encoding mast cell tryptases (TPSB1, TPSAB1) and carboxypeptidase A3 (CPA3), the beta chain of the high affinity IgE receptor (MS4A2), and prostaglandin D2 synthase (PTGDS1). We did not see a reduction in mast cell numbers in the airway mucosa of these healthy individuals, so the predominant effect of ICS appears to be on gene transcription, rather than mast cell survival, but nevertheless suggests that ICS have a potentially important dampening effect on mast cell function. In biopsies, there was also downregulation of the important mast cell chemoattractants CXCL10 and CXCL11 which promote mast cell migration to the ASM in mild steroid-naive asthma through the airway mast cell chemokine receptor CXCR3(22). In people with severe asthma using high dose ICS +/− oral corticosteroids, mast cells are not increased within the ASM (23,24), suggesting suppression of mast cell chemoattractants from ASM may remain corticosteroid-sensitive in severe disease. This is in contrast to the release of mast cell-derived proteases and autacoids which demonstrate ongoing release in severe disease(23,25).

Other highly downregulated T2-related genes included IL33, an airway epithelial cell alarmin which acts to promote initiation of T2 responses(26), CLEC10A which is a marker for alternatively activated macrophages(27), SERPINB10 which is highly upregulated on airway epithelium by IL-13 and inhibits Th2 cell apoptosis, and CCR5 whose ligands include CCL5 (RANTES), a chemoattractant for eosinophils and mast cells. Moreover PTGS1, which encodes cyclooxygenase-1 was the 7^th^ most downregulated gene in brushings. This enzyme converts arachidonate to prostaglandins and its dysregulation or inhibition is implicated in salicylate-sensitive asthma and sino-nasal eosinophilic inflammation(28). Similarly in biopsies, periostin (POSTN), an IL-13-induced epithelial gene associated with T2 high asthma(29), was also downregulated.

Taken together, the ability of ICS to downregulate this extensive set of T2-related genes implies that homeostasis in health involves a low level of tonic T2 signalling in the airway mucosa that is very sensitive to ICS. This would be consistent with the observation that IL-4- and IL-5-positive cells are present in healthy airways(30), as are mast cells and dendritic cells expressing FcεRIα(31). Constitutive T2 signalling is also evident in primary epithelial cell cultures grown at air-liquid interface, where we previously observed that STAT6-dependent genes including POSTN, CLCA1 and SERPINB2 were repressed by NFκΒ-dependent cytokine stimulation or dexamethasone(32). Thus differentiated bronchial epithelial cells have a low level of tonic STAT6 dependent signalling in the absence of exogenous IL-4 or IL-13. However, FeNO was not reduced by ICS in these healthy volunteers, and NOS2, which regulates FeNO production, was not altered by ICS therapy, suggesting tonic T2 signalling is below the threshold required for pathological FeNO generation.

The role of low-level, tonic, constitutive T2 signalling in the airways is uncertain but likely beneficial for tissue homeostasis. T2 immunity exhibits many host-protective functions, including maintaining metabolic homeostasis, suppressing excessive T1 inflammation, maintenance of barrier defence and regulation of tissue regeneration(33–36). For example, IL-33 is pleiotropic and can promote type 2 inflammation but in other contexts it can be immunoregulatory(26), and preserves epithelial integrity during influenza infection in a mouse model(37,38). Thus the effects of type 2 immunity may be context specific and affected by the cellular source and concomitant inflammatory milieu, and at steady state may promote airway epithelial barrier function. Inhibition of homeostatic tonic T2 signalling might therefore have deleterious effects which may have an impact clinically, and might explain an enhanced propensity to proteobacterial colonisation or infection, as was observed in the MEX(39) and RASP-UK(40) studies amongst participants with very low T2 biomarkers.

In addition to the inhibition of T2-driven genes, there was marked suppression of molecules involved in both protective innate immunity (e.g. CD48, CEACAM5) and adaptive immunity, with suppression of genes related to dendritic cells (CD207), T cells (e.g. CD2/3/6/8/96, TRBC1/2, TRAC), and B cell function (e,g, CD20/79, most heavy and variable light chains for IgA, IgG and IgM, JCHAIN). This potential impairment of innate, cell-mediated and antibody-dependent immunity likely explains the reproducible dose-dependent increased pneumonia risk in people with asthma and COPD who are using ICS(4). A subgroup of people with asthma also become colonised with certain fungi(41), and it is possible that this is also a consequence of the mucosal immunosuppression observed here.

An important aim of this study was to provide information on the activity of ICS in healthy airways, to remove the confounding that might occur due to disease-related changes in gene expression or inherent ICS responsiveness, which should facilitate analyses of gene expression changes in asthma. Studies of severe asthma to-date (42) have attempted to account for the effects of ICS based on gene expression changes derived from in vitro cell cultures, from an interventional study of gene expression changes in people administered ICS for chronic obstructive pulmonary disease, and from one previous intervention study which used microarrays to assess acute airway transcriptional consequences 6 hours after a single inhaled dose of budesonide (1600 µg) in 12 healthy, steroid-naïve men(43). Our findings are complementary to the previous budesonide study in that some differentially regulated genes are common to our two studies (including upregulation of FKBP5, ZBTB16, PHACTR3, TSC22D3 and downregulation of CD207, FCER1A, IL33), but there is a striking difference in the overall consequences of a single dose versus 4 weeks twice daily ICS use. The predominant effect of a single large acute dose of ICS was upregulation of genes (transactivation), with 68 genes upregulated and only 28 downregulated. Many of the upregulated genes were proinflammatory including growth factors, chemokines, chemokine receptors, cytokines, growth factors, and coagulation factors. By comparison in our chronic high dose exposure study only 53 genes were upregulated in biopsies, whilst 416 genes were downregulated (transrepression). This highlights the critical and often overlooked importance of the complex temporal dynamics of corticosteroid effects. Early during an acute infectious or traumatic challenge a surge in endogenous corticosteroids will tend to enhance protective inflammatory and procoagulant responses, but as time passes these responses will attenuate and slower but potent transrepressive effects will dominate to curtail uncontrolled inflammatory cell recruitment and activation, to prevent bystander tissue damage, and to direct resolution of tissue homeostasis. It is this latter situation which is of greatest clinical relevance to long term asthma management. Of note, comparing our data to the study by Woodruff(12), where people with mild asthma received inhaled placebo or fluticasone propionate (500 µg) twice daily for 8 weeks, there was a strong correlation between the studies for ICS-inducible genes, suggesting that in mild asthma at least, ICS-dependent transactivation is preserved.

When considering previous studies of gene expression in severe asthma, an IL-17-dependent gene signature is expressed in a subset of people, and mutually exclusive with a T2 gene signature(8,9). This IL-17-dependent signature was only seen in people using ICS, but it has been unclear whether this is disease- or treatment-related, and whether it is harmful or potentially protective. We did not see upregulation of this IL-17 signature after 4 weeks of ICS treatment in this study, thus it seems most likely a feature of disease. Similarly, there is upregulation of CEACAM family members in severe asthma, notably the IL-13-dependent gene CEACAM5(44,45), and CEACAM6 which was increased on both the epithelium and neutrophils in bronchial biopsies(44). Here we found that CEACAM6 expression did not change with ICS treatment, while CEACAM5 expression was reduced (tables E2, E4), in keeping with the inhibition of other T2-related genes. Therefore the upregulation of these CEACAMs in severe asthma appears to be a feature of the disease rather than treatment. In the recent U-BIOPRED bronchoscopy study(24), ICS-inducible genes such as FKBP5 were upregulated in severe asthma, suggesting that their participants were adherent to treatment, and importantly, that ICS transactivation appears to be preserved in severe asthma, but many pathological pathways that should be sensitive to transrepression by ICS such as T2 signalling are not responsive in a subset of patients(8,9,23).

There are some limitations to our work. Firstly the effects of ICS in healthy airways at 4 weeks, while likely representative of the steady state in long term therapy, might not be fully representative of longer term therapy. However it is not reasonable to ask healthy volunteers to take ICS for a year, and adherence would likely wane. Secondly our analyses use bulk sequencing of airway brushes and biopsies, and ICS effects are likely to be highly cell-specific, so future studies using spatial sequencing, from central and peripheral airways as well as nasal tissues and peripheral blood would be informative. For example, as we did not enumerate all of the cell types implicated in the transcriptomic analyses, we cannot conclude whether the changes in T- and B-lymphocyte related gene expression are due to transcriptional changes within lymphocytes, changes in the relative proportions of lymphocytes in the samples, or a combination of the two.

In summary, we defined genes altered directly by ICS therapy without confounding by disease. We provide evidence that IL-17-dependent signalling in asthma is disease-dependent rather than ICS-dependent, and demonstrate downregulation of canonical markers of T2 inflammation, implying that homeostasis in health involves tonic T2 signalling in the airway mucosa, which is exquisitely sensitive to ICS. There was also broad suppression of innate and adaptive immunity, in keeping with known immunosuppressive effects of corticosteroids.

## Supporting information

Supplementary methods and results

## Author contributions

All authors reviewed the data and contributed to its interpretation, edited the manuscript, and approved the final submitted version.

### Specific contributions

**EM:** Analysed the RNA seq data and drafted the manuscript

**TSCH:** Contributed to data analysis and interpretation, and drafting of the manuscript.

**MR:** Analysed the RNA seq data.

**LC and FAS:** Performed immunohistochemistry, and quantified and collated the data.

**PR:** performed and analysed DNA methylation studies

**RLC**: Designed, supervised and analysed DNA methylation studies.

**BH:** Managed the study, contributed to patient recruitment, collated the demographic data.

**CDA:** Contributed to bronchoscopy protocol design.

**JLM** and **MSK** performed DNA methylation studies.

**SS**: Analysed the RNA seq data.

**JRA:** Conceived and designed the study.

**DC:** Conceived and designed the study, analysed RNA seq data, drafted the manuscript.

**PB**: Conceived and designed the study, wrote the study protocol and ethical application, analysed the data, drafted the manuscript, performed all bronchoscopies. Chief investigator.

## Abbreviations

ASM: airway smooth muscle
FeNO: fraction of exhaled nitric oxide
FEV_1_: forced expiratory volume
ICS: inhaled corticosteroid
IQR: interquartile range
MBP: major basic protein
MC_T_: tryptase only mast cell
MC_TC_: tryptase and chymase mast cell
PG: prostaglandin
RBM: reticular basement membrane
RTU: ready-to-use
SD: standard deviation
T2: Type-2 cytokines (IL-4, IL-5, IL-13)

## Acknowledgements

The authors are grateful to all the participants who volunteered and to the clinical and research teams at all the participating centres. For the purpose of Open Access, the author has applied a CC BY public copyright licence to any Author Accepted Manuscript version arising from this submission. The views expressed are those of the authors and not necessarily those of the NHS, the NIHR or the Department of Health and Social Care. The authors vouch for the integrity and completeness of the data and the fidelity of the trial to the protocol. This study has been conducted in accordance with the Declaration of Helsinki and any applicable regulatory requirements.

## Data sharing

The data analysed and presented in this study are available from the corresponding author on reasonable request, providing the request meets local ethical and research governance criteria after publication. Patient-level data will be anonymised. The RNA Sequencing data have been deposited in the Gene Expression Omnibus (GEO) under accession number GSE242048.

