## Supplementary methods and results for "The effects of inhaled corticosteroids on healthy airways"

**Running title:** The effects of ICS on healthy airways

#### METHODS

This prospective study was approved by the East Midlands - Leicester Central Research Ethics Committee (REC)(reference 15/EM/0313) and registered at clinicaltrials.gov (NCT02476825). All participants gave written informed consent.

##### Participant population

People aged 18-65 were eligible, and all were current non-smokers with a <10 pack year smoking history. They had no prior history or clinical evidence of lower respiratory disease, and normal spirometry. Healthy volunteers with a history of rhinitis (perennial or seasonal) were required to have a $PC_{20}$ methacholine >16 mg/ml.

##### Participant characterisation

Participants underwent extensive evaluation at baseline including a full medical history, lung function testing, with bronchial challenge using methacholine where appropriate.

##### Study design

This was a randomised, open-labelled, bronchoscopy study designed to assess the effects of fluticasone propionate on airway gene expression and cellularity in healthy adult healthy volunteers without asthma. The primary endpoint was the corticosteroid-inducible gene expression pattern in healthy airways. Secondary endpoints were the relative changes from baseline in airway cellularity, DNA methylation and the microbiome following 4-weeks of inhaled fluticasone propionate treatment.

Target recruitment was 30 healthy adult subjects randomised by a blinded investigator (MR) in a 2:1 ratio to one of two study groups: i) subjects receiving fluticasone propionate 500 mcg b.i.d. via Accuhaler (Diskus) for 4 weeks (n=20), or ii) subjects who received no treatment for 4 weeks (n=10). Bronchoscopy was performed in all subjects at baseline prior to the start of the treatment period, and at the end of week 4. Genentech and Leicester laboratory support staff were blinded to treatment allocation. A control arm was included to provide a comparator for the treatment group and to assess the repeatability of the planned analyses.

To ensure there were sufficient data for analysis, if a subject withdrew before completion of the study, a further subject(s) was randomised after the first 30 randomisations until a total of 30 subjects had completed the study.

**Bronchoscopy**

Subjects underwent bronchoscopy conducted according to British Thoracic Society guidelines(E1). Mucosal biopsies and brushes were collected from 2$^{nd}$-5$^{th}$ generation bronchi under direct vision as per study procedure manual.

**Tissue processing and immunohistochemistry**

Biopsies were fixed in 4% neutral buffered formalin for 4 hours at 4°C as described(E2), then processed into paraffin wax, as per study procedure manual. Immunohistochemistry was performed in Leicester. All the laboratory procedures and processes were performed following the ISO9001-2015 Quality Management System and GCP/GLP guidelines.

3 µm tissue sections were cut and dewaxed, and stained with haematoxylin and eosin for quality control assessment, to ensure the integrity of the key tissue elements needed for immunohistochemistry staining and analysis. Further 3 µm sections were immunostained with the following mouse monoclonal primary antibodies: anti–neutrophil elastase (clone NP57, 0.42 µg/mL, Agilent Dako, UK), anti–mast cell tryptase clone AA1 (ready-to-use [RTU], Agilent Dako), anti-mast cell chymase (LS-B12242, 0.8 µg/mL, LSBio, UK), anti–eosinophil major basic protein (clone BMK13, 1 µg/mL, Monosan, UK), anti-α-smooth muscle actin (αSMA)(RTU, Dako) and appropriate isotype controls (Dako and BD Biosciences). All immunostaining steps were performed using an Autostainer Link 48 (Agilent Dako, UK) using appropriate isotype controls followed by counterstaining in Gill's haematoxylin. Two sections from 2 biopsies were stained for each parameter.

#### Assessment of immunopathology

High-throughput morphologic analysis was performed on scanned sections using the Carl Zeiss Scanner Z1 and AxioCam HRc digital camera (Carl Zeiss, Germany). ZEN desk 3.1 image analysis software was used to perform image analysis on total, ASM, epithelium and lamina propria areas. The following previously validated pathological features(E3) were evaluated as follows; i) nucleated inflammatory cells (eosinophils, neutrophils, mast cells [tryptase+ and chymase+]) were counted in the airway epithelium and lamina propria, and expressed as cells/mm$^2$ of the compartment of interest; ii) the percentage of biopsy area occupied by airway smooth muscle was measured and expressed as a percentage of the total biopsy area; iii) reticular basement membrane (RBM) thickness was expressed as the mean value of 50-point measurements approximately 20 µm apart according to the method validated by Sullivan *et al*(E4). The mean of two sections from 2 biopsies was taken for each analysis.

All pathological data were assessed by an observer blinded to the identity and treatment allocations of the participants.

#### RNA sequencing

Bronchial brushes and biopsies were collected in RNAlater as per study procedure manual. Total RNA and DNA separately were isolated from the bronchial brushes and biopsies using QIAgen AllPrep DNA/RNA/miRNA Universal Mini Kit per manufacturer's instructions. Ribosomal RNA was removed using the RiboZero Magnetic Gold kit. RNA sequencing libraries were prepared using the Illumina TruSeq Stranded Total RNA method. Libraries were single-end sequenced. The RNA Sequencing data have been deposited in the Gene Expression Omnibus (GEO) under accession number GSE242048.

#### Bisulphite conversion and DNA methylation arrays

Seven hundred fifty ng of purified genomic DNA was bisulfite converted using the EZ DNA Methylation Kit (Zymo Research) as per the manufacturer's instructions. Specific incubation conditions for the Illumina Infinium Methylation Assay were used as per the manufacturer's protocol Appendix. Samples were eluted in 12 µl of the provided elution buffer. Bisulfite-converted DNA was assessed for concentration and quality using a NanoDrop™ 8000 Spectrophotometer (Thermo Fisher Scientific), and 160 ng of the conversion product was used for genome-wide DNA methylation quantification at over 850,000 CpG sites using the Illumina Infinium HumanMethylationEPIC BeadChip array, according to the manufacturer's protocols.

**DNA methylation data quality control and normalization**

Raw data were obtained from GenomeStudio software in IDAT format. The data were analysed using R statistical software (version 4.1.0). Pre-processing and quality control were performed using *minfi* package (version 1.38.0) (E5). The 65 known quality control SNP probes were used to cluster all samples to detect anomalies within samples from the same donor. Probes were excluded from further analysis according to several criteria: first, the 65 SNP probes, second, 84,953 probes represented by less than three beads in at least 5% of samples; third, 1,721 probes that did not pass detection p value threshold ($p<0.05$) in at least 1% of the samples; fourth, 17,323 probes located on X and Y chromosomes; and finally 26,641 probes that overlap single nucleotide polymorphism and 35979 probes that cross-hybridise to multiple regions on the genome (E6). 699,474 probes remained for analysis. Functional normalisation of filtered probes was carried out using *preprocessFunnorm* function in *minfi* package. Batch effect correction was performed on normalised data to correct for chip and sample position bias using *comBat* function in *sva* package (version 3.40.0)(E7). Two values of DNA methylation were calculated, beta-values ($\beta$-values) and *M*-values. $\beta$-values are the ratio of all methylated probe intensities over total signal intensities (methylated and unmethylated) and have a range from 0 to 1. They approximately represent percent methylation. *M*-values are the logit transformation of $\beta$-values and are more statistically robust(E8). All statistical analyses were performed using *M*-values, while $\beta$-values were used for visualization and interpretability purposes.

**Differential DNA methylation analysis**

Linear regression analysis was applied to the data using the *limma* package (version 3.48.0) in R(E9). An interaction term and subject correlation were included to identify any probes differentially methylated between baseline and week 4 in individuals taking fluticasone independently to the control untreated individuals (Methylation~Timepoint*Treatment + [1 | Subject ID]).

**Expression quantitative trait methylation (eQTM) analysis**

Association of DNA methylation with differential expression was studied using expression quantitative trait methylation analysis. Twenty nine differentially expressed genes and comprehensive DNA methylation data (699474 probes resulting after probe filtering) from week 0 and week 4 fluticasone treatment samples were given as input into an eQTM linear regression model to identify significantly correlating expression-methylation pairs. Significant pairs were classified based on a Benjamini-Hochberg false discovery rate (FDR) less than 0.05.

**Microbiota Sequence Data Generation, Processing, and Analysis**

DNA isolated from bronchial brushes was sent to Diversigen for bacterial 16SV4 rRNA gene sequencing as previously described(E10,11). Briefly, PCR amplification of the 16S rRNA gene was conducted for each sample using barcoded primers targeting the 16S-V4 region. Following PCR, amplicon libraries were pooled at equimolar concentrations and purified. Pooled amplicon sequence libraries were then sequenced with a MiSeq 600 cycle v3 kit (Illumina, San Diego, CA). Following sequencing, QIIME2 v2019.7 was used to process the 16S-V4 rRNA gene sequence data as previously described(E12,13). Briefly, raw sequence data was demultiplexed, read trimming was performed to remove regions of low sequence quality, and paired-end reads were denoised, dereplicated, and chimera filtered with DADA2(E14). The remaining non-chimeric reads were then filtered to remove reads mapping to the human genome using bowtie2(E15). Taxonomy was assigned based on the V4 region of the Genome Taxonomy Database 16S rRNA gene sequence database (r202)(E16). Resulting abundance tables were rarefied to 8900 reads per sample to account for variable sequencing depth prior to calculation of α- and β-diversity metrics and samples with less than 8900 high quality reads were discarded. All statistical analyses were conducted in the R statistical environment as previously described (Team RC. R: A Language and Environment for Statistical Computing. Vienna, Austria; 2020)(E17). All microbiome sequence data is available from the European Genome-Phenome Archive (https://ega-archive.org/, EGAS00001007538).

##### Transcriptomic analysis

For analysis of RNA-seq data, sequences in fastq files (in single and pair ends) were aligned using STAR aligner (version 2.7.1a) to the human reference genome GRCh38; R package (E18)Rsubread was employed for quantification of reads assigned to genes.

Raw count pre-processing, normalisation and differential gene expression analysis was performed using R packaged DESeq2, edgeR, and limma. Gene pathways enrichment analysis was executed using R packages fgsea, clusterProfile, and ReactomePA. Heatmaps, volcano and MA plots were generated using in-home R code, ggplot2 and plotly packages(E18,19).

##### Statistical analysis

Basic summary statistical analysis was performed using GraphPad Prism version 7.03 (GraphPad Software, San Diego). Parametric and non-parametric data are presented as mean (standard error mean [SEM]) and median (interquartile range [IQR]) respectively unless otherwise stated.

#### SUPPLEMENTARY RESULTS

##### Effects of ICS on the airway microbiome

To assess the potential impact of ICS treatment on the bacterial community composition of the lower airways, bronchial brushing DNA from paired baseline and week 4 samples from treated and untreaded individuals was subjected to 16SV4 rRNA gene sequencing (16SV4-seq). Processing resulted in high quality 16SV4-seq data from paired baseline/week 4 samples from 6 untreated and 12 ICS-treated individuals. Between baseline and week 4, no difference in α-diversity was observed, regardless of treatment group. Additionally, no difference in gross bacterial community composition based on β-diversity was observed when comparing baseline and week 4 samples between treated and untreated participants. At the individual taxa level, the relative abundance of only two species was significantly changed (Benjamini-Hochberg adjusted p-value $< 0.05$) following ICS treatment. Relative abundance of *Alloprevotella tannerae* was significantly increased at week 4 in ICS-treated individuals compared to baseline while that of an unclassified *Corynebacterium* member was decreased (**Supplementary Figure E10**).

##### Effects of ICS on airway DNA methylation

Linear regression analysis identified only a single significantly differentially methylated CpG site, cg14383238 (Benjamini Hochberg p=0.013) (**Supplementary Figure E11**), suggesting global airway epithelial cell DNA methylation is minimally affected by short term fluticasone treatment. cg14383238 lies on north shore of a CpG island located in the promoter associated transcription start site of transmembrane protein 63C (TMEM63C) located on chromosome 14. This gene is associated with $Ca^{2+}$-activated cation channel activity required for kidney glomerular filtration barrier function.

Having identified 29 differentially expressed genes in response to four weeks inhaled corticosteroid treatment in bronchial epithelial cells, we sought to investigate the involvement of DNA methylation in regulating these changes. Expression quantitative trait methylation (eQTM) analysis was performed using differentially expressed genes and 699474 CpG sites in matched samples of bronchial brushing derived epithelial cells. There were 48978 significant gene-CpG correlations (Benjamini Hochberg $p<0.05$) that comprised 29 differentially expressed genes and 30389 CpG sites. Despite the correlation, there was no significant difference relative to baseline in the expression-associated CpG sites following 4 weeks of fluticasone treatment.

E12. Wagner F, Mansfield JC, Lekkerkerker AN, Wang Y, Keir M, Dash A, Butcher B, Harder B, Orozco LD, Mar JS, Chen H, Rothenberg ME. Dose escalation randomised study of efmarodocokin alfa in healthy volunteers and patients with ulcerative colitis. *Gut* 2023.

E13. Bolyen E, Rideout JR, Dillon MR, Bokulich NA, Abnet CC, Al-Ghalith GA, Alexander H, Alm EJ, Arumugam M, Asnicar F, Bai Y, Bisanz JE, Bittinger K, Brejnrod A, Brislawn CJ, Brown CT, Callahan BJ, Caraballo-Rodriguez AM, Chase J, Cope EK, Da Silva R, Diener C, Dorrestein PC, Douglas GM, Durall DM, Duvallet C, Edwardson CF, Ernst M, Estaki M, Fouquier J, Gauglitz JM, Gibbons SM, Gibson DL, Gonzalez A, Gorlick K, Guo J, Hillmann B, Holmes S, Holste H, Huttenhower C, Huttley GA, Janssen S, Jarmusch AK, Jiang L, Kaehler BD, Kang KB, Keefe CR, Keim P, Kelley ST, Knights D, Koester I, Kosciolek T, Kreps J, Langille MGI, Lee J, Ley R, Liu YX, Loftfield E, Lozupone C, Maher M, Marotz C, Martin BD, McDonald D, McIver LJ, Melnik AV, Metcalf JL, Morgan SC, Morton JT, Naimey AT, Navas-Molina JA, Nothias LF, Orchanian SB, Pearson T, Peoples SL, Petras D, Preuss ML, Pruesse E, Rasmussen LB, Rivers A, Robeson MS, 2nd, Rosenthal P, Segata N, Shaffer M, Shiffer A, Sinha R, Song SJ, Spear JR, Swafford AD, Thompson LR, Torres PJ, Trinh P, Tripathi A, Turnbaugh PJ, Ul-Hasan S, van der Hooft JJJ, Vargas F, Vazquez-Baeza Y, Vogtmann E, von Hippel M, Walters W, Wan Y, Wang M, Warren J, Weber KC, Williamson CHD, Willis AD, Xu ZZ, Zaneveld JR, Zhang Y, Zhu Q,

Knight R, Caporaso JG. Reproducible, interactive, scalable and extensible microbiome data science using QIIME 2. *Nat Biotechnol* 2019; 37: 852-857.
E14. Callahan BJ, McMurdie PJ, Rosen MJ, Han AW, Johnson AJ, Holmes SP. DADA2: High-resolution sample inference from Illumina amplicon data. *Nat Methods* 2016; 13: 581-583.
E15. Langmead B, Salzberg SL. Fast gapped-read alignment with Bowtie 2. *Nat Methods* 2012; 9: 357-359.
E16. Parks DH, Chuvochina M, Chaumeil PA, Rinke C, Mussig AJ, Hugenholtz P. A complete domain-to-species taxonomy for Bacteria and Archaea. *Nat Biotechnol* 2020; 38: 1079-1086.
E17. Mar JS, Ota N, Pokorzynski ND, Peng Y, Jaochico A, Sangaraju D, Skippington E, Lekkerkerker AN, Rothenberg ME, Tan MW, Yi T, Keir ME. IL-22 alters gut microbiota composition and function to increase aryl hydrocarbon receptor activity in mice and humans. *Microbiome* 2023; 11: 47.
E18. Foundation TR. The R Project for Statistical Computing. [cited 2023 23/05/2023]. Available from: http://www.r-project.org/index.html.
E19. Reimers M, Carey VJ. Bioconductor: an open source framework for bioinformatics and computational biology. *Methods Enzymol* 2006; 411: 119-134.
E20. Woodruff PG, Boushey HA, Dolganov GM, Barker CS, Yang YH, Donnelly S, Ellwanger A, Sidhu SS, Dao-Pick TP, Pantoja C, Erle DJ, Yamamoto KR, Fahy JV. Genome-wide profiling identifies epithelial cell genes associated with asthma and with treatment response to corticosteroids. *Proc Natl Acad Sci U S A* 2007; 104: 15858-15863.

**Supplementary Table E1.** All upregulated differentially expressed genes in bronchial brushes after 4 weeks of inhaled fluticasone (treatment group only). Genes shown are censored at FDR P < 0.05 and log(2) fold change of ±1 and ordered by P value. FDR, false discovery rate.

**Genes upregulated in bronchial brushes**

| Gene | Baseline read count | Log2 fold change | t | *P* value | FDR *P* value |
|---|---|---|---|---|---|
| PHACTR3 | 497 | 3.59 | 14.22 | 7.3E-46 | 1E-41 |
| HSD11B2 | 468 | 1.89 | 14.11 | 3.3E-45 | 3E-41 |
| MYLK3 | 476 | 1.10 | 13.99 | 1.8E-44 | 1E-40 |
| FKBP5 | 3336 | 1.88 | 11.56 | 6.7E-31 | 2E-27 |
| BTNL9 | 369 | 1.71 | 11.52 | 1.1E-30 | 3E-27 |
| GRAMD2A | 2452 | 1.15 | 10.81 | 3E-27 | 5E-24 |
| IFITM10 | 1872 | 1.55 | 10.36 | 3.9E-25 | 4E-22 |
| FAM107A | 3486 | 1.45 | 10.32 | 6E-25 | 6E-22 |
| HCAR2 | 4673 | 1.10 | 9.81 | 1E-22 | 9E-20 |
| TSC22D3 | 7099 | 1.12 | 9.28 | 1.7E-20 | 1E-17 |
| SLCO1B3 | 124 | 1.94 | 9.24 | 2.5E-20 | 2E-17 |
| SULT2B1 | 1706 | 1.38 | 9.17 | 4.7E-20 | 3E-17 |
| HIF3A | 584 | 3.18 | 9.13 | 7E-20 | 4E-17 |
| ADAMTS9 | 204 | 1.38 | 9.08 | 1.1E-19 | 6E-17 |
| CYP17A1-AS1 | 115 | 1.83 | 9.04 | 1.5E-19 | 9E-17 |
| ITGA10 | 160 | 1.41 | 8.73 | 2.5E-18 | 1E-15 |
| GP2 | 341 | 2.48 | 8.74 | 2.4E-18 | 1E-15 |
| KCNB1 | 602 | 1.03 | 8.68 | 3.8E-18 | 2E-15 |
| SYT8 | 503 | 1.12 | 8.62 | 6.6E-18 | 3E-15 |
| CD163 | 1860 | 2.45 | 8.51 | 1.8E-17 | 7E-15 |

| | | | | | |
|---|---|---|---|---|---|
| ANPEP | 2159 | 1.16 | 8.29 | 1.1E-16 | 4E-14 |
| PRODH | 5268 | 1.20 | 8.26 | 1.5E-16 | 6E-14 |
| PSCA | 23905 | 1.21 | 8.19 | 2.6E-16 | 9E-14 |
| TFCP2L1 | 3228 | 1.38 | 8.06 | 7.9E-16 | 3E-13 |
| GAS6-AS1 | 121 | 1.42 | 7.86 | 4E-15 | 1E-12 |
| GNMT | 704 | 1.28 | 7.77 | 7.9E-15 | 2E-12 |
| TAT-AS1 | 242 | 1.01 | 7.58 | 3.4E-14 | 8E-12 |
| ANGPT1 | 188 | 1.22 | 7.41 | 1.3E-13 | 3E-11 |
| ABCC3 | 882 | 1.00 | 7.23 | 4.7E-13 | 9E-11 |
| PPP1R16B | 2159 | 1.11 | 7.11 | 1.2E-12 | 2E-10 |
| KCNMA1 | 369 | 1.12 | 6.99 | 2.8E-12 | 5E-10 |
| GAS1 | 136 | 1.15 | 6.80 | 1E-11 | 1E-09 |
| LPL | 369 | 2.41 | 6.65 | 2.8E-11 | 4E-09 |
| LINC00930 | 204 | 1.11 | 6.50 | 7.9E-11 | 9E-09 |
| FLVCR2 | 173 | 1.01 | 6.41 | 1.5E-10 | 2E-08 |
| MUC21 | 134 | 1.51 | 6.28 | 3.5E-10 | 3E-08 |
| HAP1 | 132 | 1.17 | 6.21 | 5.3E-10 | 5E-08 |
| VSIG4 | 808 | 1.79 | 5.98 | 2.3E-09 | 2E-07 |
| MMP7 | 203 | 1.09 | 5.95 | 2.7E-09 | 2E-07 |
| ADAMTSL4 | 232 | 1.07 | 5.80 | 6.5E-09 | 4E-07 |
| CPM | 202 | 1.30 | 5.73 | 1E-08 | 6E-07 |
| ERVS71-1 | 1059 | 1.51 | 5.72 | 1E-08 | 6E-07 |
| ZBTB16 | 362 | 1.88 | 5.70 | 1.2E-08 | 7E-07 |
| MMP19 | 193 | 2.05 | 5.66 | 1.5E-08 | 8E-07 |
| GDNF-AS1 | 119 | 1.02 | 5.58 | 2.4E-08 | 1E-06 |
| MTUS2 | 198 | 1.03 | 5.31 | 1.1E-07 | 5E-06 |
| MS4A4A | 231 | 1.19 | 5.14 | 2.7E-07 | 1E-05 |
| CYP4A11 | 175 | 1.34 | 5.10 | 3.4E-07 | 1E-05 |

| | | | | | |
|---|---|---|---|---|---|
| GLDN | 248 | 1.17 | 5.10 | 3.4E-07 | 1E-05 |
| SLC11A1 | 627 | 1.32 | 4.73 | 2.3E-06 | 6E-05 |
| AOC3 | 320 | 1.67 | 4.72 | 2.4E-06 | 6E-05 |
| MME | 291 | 1.44 | 4.65 | 3.4E-06 | 8E-05 |
| LILRA6 | 117 | 1.08 | 4.08 | 4.4E-05 | 0.0007 |
| TREM1 | 436 | 1.26 | 3.90 | 9.5E-05 | 0.0013 |
| FABP4 | 1683 | 1.49 | 3.86 | 0.00011 | 0.0014 |
| TRNA | 293 | 1.30 | 3.80 | 0.00015 | 0.0018 |
| IL1R2 | 259 | 1.45 | 3.73 | 0.00019 | 0.0022 |
| SERPINA3 | 417 | 1.77 | 3.71 | 0.00021 | 0.0023 |
| KRT6A | 1428 | 1.25 | 3.48 | 0.00049 | 0.0046 |
| CLEC4E | 156 | 1.34 | 3.29 | 0.00101 | 0.0081 |
| ELOA3BP | 160 | 1.01 | 3.28 | 0.00104 | 0.0083 |
| MARCO | 1414 | 1.26 | 3.28 | 0.00104 | 0.0083 |
| MSR1 | 1008 | 1.06 | 3.18 | 0.00147 | 0.01 |
| TRNE | 9170 | 1.08 | 3.17 | 0.00151 | 0.01 |
| PCOLCE2 | 105 | 1.04 | 3.12 | 0.00184 | 0.01 |
| MRC1 | 381 | 1.06 | 3.11 | 0.00189 | 0.01 |
| MCEMP1 | 186 | 1.01 | 2.95 | 0.00313 | 0.02 |
| RBP4 | 164 | 1.18 | 2.86 | 0.00426 | 0.02 |
| APOC1 | 1804 | 1.05 | 2.75 | 0.00597 | 0.03 |
| CXCL13 | 101 | 1.86 | 2.64 | 0.00838 | 0.04 |
| APOC1P1 | 124 | 1.16 | 2.63 | 0.00866 | 0.041 |
| SAA1 | 23140 | 1.34 | 2.58 | 0.00992 | 0.045 |

**Supplementary table E2.** All downregulated differentially expressed genes in bronchial brushes after 4 weeks of inhaled fluticasone (treatment group only). Genes shown are censored at FDR P < 0.05 and log(2) fold change of ±1 and ordered by P value. FDR, false discovery rate.

**Genes downregulated in bronchial brushes**

| **Gene** | **Baseline read count** | **Log2 fold change** | **t** | ***P* value** | **FDR *P* value** |
|---|---|---|---|---|---|
| FCER1A | 144 | -2.78 | -10.91 | 1.019E-27 | 2.15E-24 |
| ZNF683 (Hobit) | 136 | -3.33 | -10.66 | 1.544E-26 | 2.31E-23 |
| CD207 (Langerin) | 169 | -3.78 | -10.63 | 2.193E-26 | 3.05E-23 |
| IL33 | 3885 | -1.20 | -10.43 | 1.792E-25 | 2.18E-22 |
| CLEC4F | 102 | -2.18 | -10.35 | 4.237E-25 | 4.59E-22 |
| CCL5 (RANTES) | 1133 | -2.39 | -8.74 | 2.232E-18 | 1.21E-15 |
| PTGS1 | 129 | -1.19 | -8.64 | 5.515E-18 | 2.5E-15 |
| HEPACAM2 | 450 | -1.00 | -8.59 | 8.478E-18 | 3.67E-15 |
| TARP | 100 | -2.50 | -8.51 | 1.724E-17 | 7.3E-15 |
| CD96 | 253 | -1.62 | -7.85 | 4.091E-15 | 1.31E-12 |
| ABCB11 | 147 | -1.32 | -7.52 | 5.653E-14 | 1.33E-11 |
| UCN3 | 239 | -1.05 | -7.30 | 2.792E-13 | 5.67E-11 |
| TRBC1 | 353 | -2.33 | -7.04 | 1.95E-12 | 3.39E-10 |
| CX3CR1 | 102 | -1.49 | -7.04 | 1.969E-12 | 3.4E-10 |
| TRBC2 | 463 | -2.41 | -6.92 | 4.595E-12 | 7.16E-10 |
| CD3E | 375 | -1.55 | -6.87 | 6.523E-12 | 9.48E-10 |
| PTPN22 | 165 | -1.26 | -6.80 | 1.017E-11 | 1.43E-09 |
| TRAC | 539 | -1.92 | -6.67 | 2.559E-11 | 3.37E-09 |
| IGF2BP3 | 140 | -1.29 | -6.66 | 2.77E-11 | 3.6E-09 |
| KLRB1 | 131 | -1.69 | -6.65 | 2.935E-11 | 3.76E-09 |

| | | | | | |
|---|---|---|---|---|---|
| TRAF3IP3 | 231 | -1.01 | -6.57 | 5.098E-11 | 6.37E-09 |
| CPA3 | 228 | -2.26 | -6.55 | 5.716E-11 | 7E-09 |
| GIMAP7 | 143 | -1.71 | -6.53 | 6.429E-11 | 7.68E-09 |
| CD2 | 566 | -1.87 | -6.45 | 1.141E-10 | 1.23E-08 |
| CLEC10A | 135 | -1.71 | -6.38 | 1.776E-10 | 1.82E-08 |
| SERPINB10 | 287 | -1.18 | -6.38 | 1.797E-10 | 1.82E-08 |
| SCML4 | 123 | -1.09 | -6.30 | 2.994E-10 | 2.86E-08 |
| TMC8 | 344 | -1.00 | -6.22 | 4.86E-10 | 4.36E-08 |
| CTSW | 148 | -1.42 | -6.18 | 6.362E-10 | 5.46E-08 |
| KLRC1 | 127 | -1.34 | -6.12 | 9.415E-10 | 7.84E-08 |
| RASAL3 | 240 | -1.06 | -6.11 | 9.794E-10 | 8.05E-08 |
| MUC13 | 4313 | -1.67 | -6.10 | 1.052E-09 | 8.58E-08 |
| HSPA7 | 253 | -1.30 | -6.06 | 1.334E-09 | 1.05E-07 |
| GIMAP6 | 201 | -1.55 | -6.02 | 1.756E-09 | 1.33E-07 |
| LCK | 324 | -1.15 | -5.99 | 2.16E-09 | 1.57E-07 |
| SPOCK2 | 418 | -1.29 | -5.77 | 7.949E-09 | 4.86E-07 |
| CD8A | 516 | -1.55 | -5.71 | 1.11E-08 | 6.4E-07 |
| CD8B | 171 | -1.26 | -5.67 | 1.418E-08 | 7.98E-07 |
| PLG | 110 | -1.08 | -5.62 | 1.912E-08 | 1.05E-06 |
| ITGA1 | 273 | -1.17 | -5.61 | 2.031E-08 | 1.11E-06 |
| ITK | 119 | -1.48 | -5.59 | 2.228E-08 | 1.2E-06 |
| ITLN1 | 197 | -1.93 | -5.52 | 3.433E-08 | 1.71E-06 |
| GFI1 | 133 | -1.06 | -5.52 | 3.478E-08 | 1.73E-06 |
| CD247 | 132 | -1.56 | -5.32 | 1.042E-07 | 4.61E-06 |
| CD3D | 283 | -1.66 | -5.31 | 1.1E-07 | 4.83E-06 |
| RGS7BP | 172 | -1.06 | -5.25 | 1.505E-07 | 6.32E-06 |
| KLRC2 | 106 | -1.23 | -5.20 | 2.006E-07 | 7.95E-06 |
| HLA-DQB2 | 798 | -1.01 | -5.16 | 2.481E-07 | 9.52E-06 |

| | | | | | |
|---|---|---|---|---|---|
| GLYATL2 | 190 | -1.42 | -5.06 | 4.117E-07 | 1.46E-05 |
| IKZF3 | 405 | -1.35 | -5.05 | 4.425E-07 | 1.56E-05 |
| CXCR6 | 396 | -1.65 | -5.03 | 4.95E-07 | 1.72E-05 |
| GIMAP8 | 116 | -1.21 | -5.02 | 5.168E-07 | 1.78E-05 |
| IGKC | 400 | -2.29 | -4.91 | 8.899E-07 | 2.79E-05 |
| IGHG2 | 175 | -2.35 | -4.72 | 2.315E-06 | 6.23E-05 |
| GVINP1 | 212 | -1.01 | -4.66 | 3.152E-06 | 8E-05 |
| CCR5 | 295 | -1.42 | -4.61 | 4.078E-06 | 9.97E-05 |
| LTB | 103 | -1.98 | -4.60 | 4.236E-06 | 0.0001 |
| MGAM | 455 | -1.40 | -4.60 | 4.249E-06 | 0.0001 |
| TPSAB1 | 442 | -1.65 | -4.60 | 4.31E-06 | 0.0001 |
| TPSB2 | 504 | -1.66 | -4.57 | 4.81E-06 | 0.0001 |
| IGHG3 | 144 | -2.34 | -4.56 | 5.064E-06 | 0.0001 |
| IGHA1 | 502 | -2.01 | -4.44 | 8.875E-06 | 0.0002 |
| MYO7A | 120 | -1.00 | -4.41 | 1.014E-05 | 0.0002 |
| IGHG1 | 227 | -2.24 | -4.37 | 1.242E-05 | 0.0003 |
| IGLC2 | 269 | -2.33 | -4.35 | 1.335E-05 | 0.0003 |
| ADAM19 | 191 | -2.03 | -4.34 | 1.42E-05 | 0.0003 |
| PTPN7 | 248 | -1.32 | -4.29 | 1.787E-05 | 0.0003 |
| ENPP2 | 115 | -1.09 | -4.05 | 5.074E-05 | 0.0008 |
| GPR18 | 137 | -1.50 | -4.05 | 5.228E-05 | 0.0008 |
| NKG7 | 192 | -1.66 | -3.92 | 8.676E-05 | 0.001 |
| HDAC9 | 510 | -1.20 | -3.73 | 0.0002 | 0.002 |
| GZMH | 137 | -1.51 | -3.67 | 0.0002 | 0.003 |
| UBD | 2630 | -1.40 | -3.62 | 0.0003 | 0.003 |
| CD48 | 271 | -1.14 | -3.57 | 0.0004 | 0.004 |
| IGHA2 | 201 | -1.98 | -3.54 | 0.0004 | 0.004 |
| IGLC3 | 216 | -2.28 | -3.49 | 0.0005 | 0.005 |

| | | | | | |
|---|---|---|---|---|---|
| MUC2 | 2840 | -1.12 | -3.48 | 0.0005 | 0.005 |
| FGL2 | 781 | -1.10 | -3.38 | 0.0007 | 0.006 |
| CD7 | 187 | -1.23 | -3.36 | 0.0008 | 0.007 |
| CEACAM5 | 3288 | -1.24 | -3.36 | 0.0008 | 0.007 |
| SLC5A8 | 1538 | -1.01 | -3.04 | 0.002 | 0.02 |
| IL32 | 581 | -1.09 | -2.85 | 0.004 | 0.02 |

**Supplementary table E3**. All upregulated differentially expressed genes in bronchial biopsies after 4 weeks of inhaled fluticasone (treatment group only). Genes shown are censored at FDR P < 0.05 and log(2) fold change of ±1 and ordered by P value. FDR, false discovery rate.

**Genes upregulated in bronchial biopsies**

| **Gene** | **Baseline read count** | **Log2 fold change** | **t** | ***P* value** | **FDR *P* value** |
|---|---|---|---|---|---|
| PHACTR3 | 325 | 3.28 | 13.65 | 2.082E-42 | 4.287E-38 |
| HSD11B2 | 392 | 1.81 | 13.37 | 9.595E-41 | 9.879E-37 |
| GP2 | 286 | 3.12 | 11.67 | 1.906E-31 | 9.81E-28 |
| SYT8 | 470 | 1.13 | 10.52 | 6.904E-26 | 2.37E-22 |
| HIF3A | 990 | 2.67 | 10.21 | 1.818E-24 | 5.347E-21 |
| FKBP5 | 5029 | 1.67 | 10.00 | 1.526E-23 | 3.142E-20 |
| FAM107A | 3657 | 1.60 | 9.22 | 2.932E-20 | 4.024E-17 |
| MYLK3 | 490 | 1.47 | 9.16 | 5.081E-20 | 6.54E-17 |
| TFCP2L1 | 4849 | 1.40 | 9.06 | 1.357E-19 | 1.643E-16 |
| SULT2B1 | 1199 | 1.44 | 8.73 | 2.441E-18 | 2.011E-15 |
| GRAMD2A | 2309 | 1.38 | 8.70 | 3.398E-18 | 2.691E-15 |
| GMPR | 694 | 1.09 | 8.61 | 7.136E-18 | 4.592E-15 |
| HCAR3 | 1622 | 1.10 | 8.40 | 4.361E-17 | 2.566E-14 |
| PSCA | 13008 | 1.87 | 8.35 | 7.063E-17 | 3.828E-14 |
| SLCO1B3 | 111 | 2.27 | 8.31 | 9.972E-17 | 5.106E-14 |
| HCAR2 | 3361 | 1.22 | 8.20 | 2.505E-16 | 1.146E-13 |
| ATP6V1C2 | 1007 | 1.24 | 8.03 | 9.549E-16 | 4.013E-13 |
| PRODH | 4754 | 1.10 | 7.94 | 2.075E-15 | 8.217E-13 |
| TSC22D3 | 8967 | 1.14 | 7.84 | 4.357E-15 | 1.547E-12 |
| PDK4 | 2453 | 1.27 | 7.44 | 1.031E-13 | 2.869E-11 |

| | | | | | |
|---|---|---|---|---|---|
| IFITM10 | 1816 | 1.23 | 7.40 | 1.407E-13 | 3.713E-11 |
| ANPEP | 1707 | 1.02 | 7.31 | 2.592E-13 | 6.28E-11 |
| KCNB1 | 662 | 1.28 | 6.97 | 3.25E-12 | 6.626E-10 |
| ERVS71-1 | 810 | 2.02 | 6.70 | 2.045E-11 | 3.29E-09 |
| NPTX2 | 109 | 1.63 | 6.70 | 2.15E-11 | 3.405E-09 |
| PAQR4 | 943 | 1.02 | 6.59 | 4.3E-11 | 5.982E-09 |
| PKIB | 2384 | 1.21 | 6.34 | 2.364E-10 | 2.735E-08 |
| GNMT | 548 | 1.10 | 5.97 | 2.427E-09 | 1.968E-07 |
| TAT-AS1 | 146 | 1.10 | 5.82 | 5.916E-09 | 4.094E-07 |
| AACSP1 | 109 | 1.29 | 5.48 | 4.197E-08 | 2.199E-06 |
| SEC14L5 | 584 | 1.08 | 5.38 | 7.27E-08 | 3.506E-06 |
| GALNT15 | 461 | 2.29 | 4.55 | 5.267E-06 | 0.0001 |
| CLDN8 | 1164 | 1.16 | 4.52 | 6.297E-06 | 0.0002 |
| MTUS2 | 196 | 1.30 | 4.48 | 7.342E-06 | 0.0002 |
| FOXN4 | 222 | 1.03 | 4.47 | 7.923E-06 | 0.0002 |
| CDC20B | 903 | 1.15 | 4.45 | 8.653E-06 | 0.0002 |
| LINC00964 | 534 | 1.13 | 4.30 | 1.738E-05 | 0.0004 |
| KLHDC8A | 405 | 1.21 | 4.25 | 2.108E-05 | 0.0005 |
| MUC21 | 117 | 1.36 | 4.24 | 2.23E-05 | 0.0005 |
| SLC6A1 | 179 | 1.30 | 3.80 | 0.0001423 | 0.0023 |
| ZBTB16 | 759 | 1.11 | 3.79 | 0.0001514 | 0.002 |
| SERPINA3 | 1487 | 1.41 | 3.68 | 0.0002316 | 0.003 |
| APOD | 6137 | 1.09 | 3.64 | 0.0002693 | 0.004 |
| CD163 | 887 | 1.06 | 3.50 | 0.0004666 | 0.006 |
| PRH2 | 3255 | 1.52 | 3.15 | 0.0016383 | 0.02 |
| H1-3 | 144 | 1.02 | 3.05 | 0.0022815 | 0.02 |
| PRB3 | 26511 | 1.47 | 2.98 | 0.0028657 | 0.03 |
| FOSB | 812 | 1.46 | 2.97 | 0.0029993 | 0.03 |

| | | | | | |
|---|---|---|---|---|---|
| SAA1 | 8478 | 1.09 | 2.96 | 0.0030546 | 0.03 |
| COMP | 185 | 1.18 | 2.91 | 0.0036188 | 0.03 |
| PRB1 | 6653 | 1.55 | 2.88 | 0.0039479 | 0.03 |
| PRB4 | 15992 | 1.59 | 2.85 | 0.0043256 | 0.03 |
| PRB2 | 6775 | 1.53 | 2.85 | 0.0043641 | 0.03 |

**Supplementary table E4.** All downregulated differentially expressed genes in bronchial biopsies after 4 weeks of inhaled fluticasone (treatment group only). Genes shown are censored at FDR P < 0.05 and log(2) fold change of ±1 and ordered by P value. FDR, false discovery rate.

**Genes downregulated in bronchial biopsies**

| **Gene** | **Baseline read count** | **Log2 fold change** | **t** | ***P* value** | **FDR *P* value** |
|---|---|---|---|---|---|
| FCER1A | 228 | -2.25 | -11.91 | 1.01E-32 | 6.93E-29 |
| ZNF683 (Hobit) | 131 | -3.06 | -10.10 | 5.78E-24 | 1.32E-20 |
| CD207 (Langerin) | 144 | -2.90 | -9.48 | 2.47E-21 | 4.24E-18 |
| HLA-DQB2 | 640 | -1.51 | -9.47 | 2.92E-21 | 4.63E-18 |
| TNFSF15 | 212 | -1.56 | -9.29 | 1.58E-20 | 2.32E-17 |
| CCL5 (RANTES) | 1233 | -2.15 | -8.98 | 2.72E-19 | 2.67E-16 |
| IL33 | 6985 | -1.24 | -8.66 | 4.85E-18 | 3.70E-15 |
| P4HA3 | 105 | -1.72 | -8.64 | 5.40E-18 | 3.83E-15 |
| DPP4 | 481 | -1.02 | -8.62 | 6.91E-18 | 4.59E-15 |
| CD96 | 399 | -1.64 | -8.36 | 6.27E-17 | 3.49E-14 |
| THEMIS | 146 | -1.71 | -8.36 | 6.14E-17 | 3.49E-14 |
| CLEC10A | 199 | -2.20 | -8.33 | 8.31E-17 | 4.39E-14 |
| CD8A | 611 | -1.52 | -8.30 | 1.02E-16 | 5.11E-14 |
| CPVL | 624 | -1.16 | -8.25 | 1.61E-16 | 7.91E-14 |
| TRGC2 | 103 | -1.98 | -8.23 | 1.93E-16 | 9.23E-14 |
| CD2 | 732 | -1.87 | -8.22 | 2.05E-16 | 9.60E-14 |
| TRBC2 | 741 | -2.07 | -8.08 | 6.67E-16 | 2.98E-13 |
| TRBC1 | 540 | -2.07 | -8.05 | 8.33E-16 | 3.65E-13 |
| TRAC | 764 | -1.84 | -8.02 | 1.09E-15 | 4.48E-13 |
| DOCK10 | 713 | -1.55 | -7.90 | 2.77E-15 | 1.07E-12 |

| | | | | | |
|---|---|---|---|---|---|
| CCR5 | 426 | -1.91 | -7.90 | 2.90E-15 | 1.11E-12 |
| SERPINB10 | 350 | -1.42 | -7.88 | 3.41E-15 | 1.25E-12 |
| CD3G | 114 | -1.74 | -7.82 | 5.49E-15 | 1.91E-12 |
| UBASH3A | 123 | -1.92 | -7.78 | 7.42E-15 | 2.55E-12 |
| NA | 131 | -3.10 | -7.71 | 1.29E-14 | 4.27E-12 |
| P2RY10 | 115 | -1.90 | -7.64 | 2.15E-14 | 6.71E-12 |
| SPN | 381 | -1.55 | -7.61 | 2.65E-14 | 8.01E-12 |
| PIK3CG | 263 | -1.59 | -7.34 | 2.17E-13 | 5.45E-11 |
| PTPN22 | 243 | -1.52 | -7.33 | 2.32E-13 | 5.76E-11 |
| RUFY4 | 117 | -1.54 | -7.28 | 3.44E-13 | 8.04E-11 |
| NAPSB | 333 | -1.52 | -7.20 | 5.94E-13 | 1.38E-10 |
| GPRIN3 | 376 | -1.60 | -7.18 | 7.17E-13 | 1.62E-10 |
| PTPN7 | 312 | -1.64 | -7.12 | 1.07E-12 | 2.36E-10 |
| GFI1 | 177 | -1.23 | -7.11 | 1.14E-12 | 2.46E-10 |
| DNASE1L3 | 156 | -1.79 | -7.10 | 1.29E-12 | 2.76E-10 |
| CCR6 | 142 | -1.27 | -7.09 | 1.30E-12 | 2.76E-10 |
| TARP | 112 | -1.90 | -7.07 | 1.52E-12 | 3.18E-10 |
| IGHV1-46 | 241 | -2.93 | -7.00 | 2.62E-12 | 5.40E-10 |
| IGLV1-50 | 195 | -2.49 | -6.96 | 3.38E-12 | 6.83E-10 |
| ITGB7 | 427 | -1.12 | -6.85 | 7.46E-12 | 1.45E-09 |
| CPA3 | 913 | -1.98 | -6.84 | 7.90E-12 | 1.51E-09 |
| CD8B | 206 | -1.38 | -6.79 | 1.09E-11 | 1.99E-09 |
| LCK | 441 | -1.38 | -6.78 | 1.17E-11 | 2.11E-09 |
| KLRC2 | 101 | -1.29 | -6.77 | 1.26E-11 | 2.24E-09 |
| FPR3 | 291 | -1.65 | -6.77 | 1.28E-11 | 2.26E-09 |
| FCGR2B | 224 | -1.57 | -6.76 | 1.34E-11 | 2.31E-09 |
| ELK2AP | 1234 | -3.29 | -6.76 | 1.33E-11 | 2.31E-09 |
| HLA-DPB2 | 156 | -1.28 | -6.75 | 1.45E-11 | 2.49E-09 |

| | | | | | |
|---|---|---|---|---|---|
| ZNF831 | 150 | -1.54 | -6.74 | 1.56E-11 | 2.66E-09 |
| LY9 | 150 | -1.23 | -6.74 | 1.58E-11 | 2.67E-09 |
| TLR7 | 127 | -1.47 | -6.72 | 1.83E-11 | 2.99E-09 |
| COL6A5 | 136 | -3.00 | -6.68 | 2.45E-11 | 3.83E-09 |
| APLNR | 328 | -3.17 | -6.67 | 2.49E-11 | 3.86E-09 |
| IGKV1D-8 | 209 | -2.99 | -6.67 | 2.54E-11 | 3.87E-09 |
| CD3D | 291 | -1.64 | -6.66 | 2.74E-11 | 4.15E-09 |
| CXCR6 | 432 | -1.71 | -6.66 | 2.82E-11 | 4.21E-09 |
| IGHG4 | 4984 | -3.20 | -6.65 | 2.85E-11 | 4.22E-09 |
| IGHV3OR16-8 | 106 | -3.11 | -6.65 | 3.03E-11 | 4.43E-09 |
| CX3CR1 | 241 | -1.30 | -6.60 | 4.22E-11 | 5.96E-09 |
| HLA-DPB1 | 5797 | -1.25 | -6.58 | 4.73E-11 | 6.46E-09 |
| IKZF3 | 711 | -1.39 | -6.56 | 5.45E-11 | 7.33E-09 |
| IGKV1D-17 | 284 | -3.12 | -6.55 | 5.83E-11 | 7.79E-09 |
| IGKV2-40 | 197 | -2.98 | -6.54 | 6.18E-11 | 8.21E-09 |
| LOC642131 | 282 | -2.41 | -6.46 | 1.04E-10 | 1.32E-08 |
| IGHV4-4 | 204 | -2.84 | -6.46 | 1.05E-10 | 1.33E-08 |
| IGHV3-48 | 573 | -2.85 | -6.44 | 1.19E-10 | 1.50E-08 |
| LOC102724760 | 122 | -2.23 | -6.43 | 1.25E-10 | 1.55E-08 |
| HDAC9 | 686 | -1.14 | -6.42 | 1.39E-10 | 1.72E-08 |
| IGHV3-11 | 470 | -3.00 | -6.41 | 1.47E-10 | 1.81E-08 |
| HLA-DQA2 | 1904 | -1.36 | -6.40 | 1.51E-10 | 1.85E-08 |
| IGKV4-1 | 807 | -3.16 | -6.39 | 1.67E-10 | 2.03E-08 |
| IGKV1-8 | 247 | -3.17 | -6.39 | 1.71E-10 | 2.04E-08 |
| CCR2 | 290 | -1.84 | -6.39 | 1.70E-10 | 2.04E-08 |
| AKAP5 | 108 | -1.01 | -6.38 | 1.74E-10 | 2.07E-08 |
| LAX1 | 112 | -1.59 | -6.34 | 2.26E-10 | 2.63E-08 |
| NA | 120 | -3.05 | -6.33 | 2.44E-10 | 2.80E-08 |

| | | | | | |
|---|---|---|---|---|---|
| IGKV1-16 | 430 | -3.31 | -6.33 | 2.52E-10 | 2.88E-08 |
| IGHJ6 | 119 | -2.63 | -6.32 | 2.61E-10 | 2.94E-08 |
| SAMSN1 | 213 | -1.51 | -6.32 | 2.64E-10 | 2.96E-08 |
| ITGA4 | 748 | -1.02 | -6.28 | 3.38E-10 | 3.67E-08 |
| IGHV3-35 | 105 | -2.78 | -6.25 | 4.02E-10 | 4.29E-08 |
| MYO7A | 150 | -1.33 | -6.25 | 4.11E-10 | 4.37E-08 |
| SOSTDC1 | 219 | -1.12 | -6.25 | 4.22E-10 | 4.46E-08 |
| SLAMF1 | 162 | -1.78 | -6.24 | 4.33E-10 | 4.55E-08 |
| HLA-DQA1 | 3663 | -1.31 | -6.23 | 4.74E-10 | 4.93E-08 |
| HLA-DQB1 | 4235 | -1.04 | -6.22 | 5.10E-10 | 5.22E-08 |
| CD3E | 601 | -1.38 | -6.21 | 5.34E-10 | 5.45E-08 |
| IGLV1-40 | 629 | -2.83 | -6.21 | 5.41E-10 | 5.49E-08 |
| IGHV4-55 | 245 | -2.58 | -6.20 | 5.79E-10 | 5.81E-08 |
| LOC102724971 | 328 | -2.49 | -6.19 | 6.07E-10 | 6.00E-08 |
| MPEG1 | 1046 | -1.51 | -6.19 | 6.15E-10 | 6.06E-08 |
| KEL | 124 | -1.45 | -6.18 | 6.22E-10 | 6.10E-08 |
| IGHV4-31 | 178 | -2.80 | -6.18 | 6.51E-10 | 6.35E-08 |
| IGHV3-71 | 152 | -2.82 | -6.17 | 7.01E-10 | 6.75E-08 |
| SCML4 | 246 | -1.30 | -6.16 | 7.14E-10 | 6.83E-08 |
| SPOCK2 | 833 | -1.32 | -6.16 | 7.29E-10 | 6.95E-08 |
| IGKV1-6 | 331 | -3.00 | -6.15 | 7.53E-10 | 7.14E-08 |
| KLRC1 | 124 | -1.20 | -6.14 | 8.12E-10 | 7.63E-08 |
| IGKV3-7 | 256 | -2.97 | -6.11 | 9.70E-10 | 9.03E-08 |
| IGHV1-18 | 424 | -3.04 | -6.11 | 1.02E-09 | 9.44E-08 |
| IGHG3 | 11442 | -3.13 | -6.10 | 1.04E-09 | 9.57E-08 |
| SASH3 | 585 | -1.44 | -6.08 | 1.19E-09 | 1.09E-07 |
| AOAH | 400 | -1.28 | -6.07 | 1.32E-09 | 1.20E-07 |
| KCNA3 | 252 | -1.80 | -6.06 | 1.33E-09 | 1.20E-07 |

| | | | | | |
|---|---|---|---|---|---|
| CD226 | 204 | -1.21 | -6.06 | 1.33E-09 | 1.20E-07 |
| THY1 | 274 | -2.18 | -6.06 | 1.39E-09 | 1.24E-07 |
| IGKV3D-7 | 660 | -2.86 | -6.06 | 1.40E-09 | 1.24E-07 |
| GRAP2 | 236 | -1.54 | -6.05 | 1.47E-09 | 1.29E-07 |
| MS4A2 | 264 | -2.20 | -6.04 | 1.52E-09 | 1.34E-07 |
| IGKV1-13 | 363 | -2.98 | -6.02 | 1.75E-09 | 1.50E-07 |
| LCP2 | 550 | -1.32 | -6.02 | 1.76E-09 | 1.51E-07 |
| IGHV3-65 | 128 | -2.87 | -6.01 | 1.80E-09 | 1.53E-07 |
| IGKV3-15 | 1137 | -3.15 | -6.01 | 1.83E-09 | 1.54E-07 |
| KLRB1 | 226 | -1.50 | -6.00 | 2.01E-09 | 1.67E-07 |
| SLC5A7 | 236 | -1.20 | -5.98 | 2.17E-09 | 1.79E-07 |
| IGHV3-53 | 279 | -2.86 | -5.96 | 2.51E-09 | 2.02E-07 |
| IGKV1-17 | 498 | -3.15 | -5.96 | 2.59E-09 | 2.06E-07 |
| IGKV2D-40 | 196 | -3.05 | -5.96 | 2.58E-09 | 2.06E-07 |
| ZAP70 | 461 | -1.09 | -5.95 | 2.65E-09 | 2.10E-07 |
| JCHAIN | 6687 | -2.83 | -5.95 | 2.72E-09 | 2.14E-07 |
| FAM78A | 254 | -1.20 | -5.94 | 2.82E-09 | 2.20E-07 |
| IGHV3-72 | 126 | -2.97 | -5.92 | 3.20E-09 | 2.47E-07 |
| GPR171 | 112 | -1.39 | -5.92 | 3.23E-09 | 2.48E-07 |
| IGHV3-41 | 107 | -2.82 | -5.90 | 3.54E-09 | 2.68E-07 |
| IGKV1D-39 | 1300 | -3.05 | -5.89 | 3.97E-09 | 2.98E-07 |
| TESPA1 | 139 | -1.81 | -5.87 | 4.30E-09 | 3.18E-07 |
| IL10RA | 654 | -1.26 | -5.87 | 4.34E-09 | 3.19E-07 |
| CD101 | 120 | -1.06 | -5.87 | 4.49E-09 | 3.25E-07 |
| IGHV3-43 | 129 | -2.72 | -5.86 | 4.53E-09 | 3.27E-07 |
| RASAL3 | 411 | -1.32 | -5.86 | 4.55E-09 | 3.27E-07 |
| IGHG2 | 8611 | -3.12 | -5.86 | 4.71E-09 | 3.34E-07 |
| IGKV2D-29 | 109 | -2.79 | -5.86 | 4.69E-09 | 3.34E-07 |

| | | | | | |
|---|---|---|---|---|---|
| IGLC5 | 323 | -2.34 | -5.86 | 4.70E-09 | 3.34E-07 |
| CD6 | 416 | -1.03 | -5.85 | 4.96E-09 | 3.51E-07 |
| IGKV3D-15 | 876 | -3.07 | -5.84 | 5.10E-09 | 3.59E-07 |
| IL12RB1 | 133 | -1.36 | -5.83 | 5.51E-09 | 3.85E-07 |
| CYSLTR1 | 265 | -1.27 | -5.82 | 6.06E-09 | 4.16E-07 |
| IGHV3-16 | 120 | -2.86 | -5.81 | 6.11E-09 | 4.18E-07 |
| IGHV3-21 | 634 | -2.84 | -5.81 | 6.17E-09 | 4.18E-07 |
| CD5 | 222 | -1.29 | -5.80 | 6.67E-09 | 4.44E-07 |
| IGKJ1 | 167 | -2.66 | -5.80 | 6.65E-09 | 4.44E-07 |
| GPR18 | 146 | -1.62 | -5.79 | 7.19E-09 | 4.74E-07 |
| IGHJ5 | 133 | -2.75 | -5.79 | 7.24E-09 | 4.76E-07 |
| IGKV1-9 | 517 | -2.87 | -5.78 | 7.61E-09 | 4.96E-07 |
| SEMA7A | 113 | -1.33 | -5.77 | 7.74E-09 | 5.02E-07 |
| IGHV1-69 | 436 | -2.82 | -5.77 | 8.09E-09 | 5.22E-07 |
| IRF8 | 423 | -1.53 | -5.76 | 8.40E-09 | 5.37E-07 |
| IGHV3-66 | 270 | -2.86 | -5.75 | 8.77E-09 | 5.57E-07 |
| IGHJ4 | 276 | -2.81 | -5.74 | 9.32E-09 | 5.89E-07 |
| NCKAP1L | 586 | -1.27 | -5.74 | 9.60E-09 | 6.02E-07 |
| IGHV2-70 | 103 | -2.79 | -5.73 | 9.81E-09 | 6.10E-07 |
| PTPRC | 2347 | -1.50 | -5.72 | 1.07E-08 | 6.62E-07 |
| IGHG1 | 20412 | -3.19 | -5.72 | 1.08E-08 | 6.65E-07 |
| IGLV3-10 | 287 | -3.02 | -5.71 | 1.11E-08 | 6.77E-07 |
| IGHV3-22 | 137 | -3.04 | -5.71 | 1.14E-08 | 6.94E-07 |
| IL2RG | 895 | -1.36 | -5.69 | 1.24E-08 | 7.51E-07 |
| IGHV3-73 | 145 | -2.50 | -5.68 | 1.37E-08 | 8.23E-07 |
| IGKV1-27 | 228 | -2.70 | -5.67 | 1.39E-08 | 8.28E-07 |
| IGKJ4 | 151 | -2.94 | -5.67 | 1.40E-08 | 8.29E-07 |
| GVINP1 | 471 | -1.20 | -5.66 | 1.49E-08 | 8.77E-07 |

| | | | | | |
|---|---|---|---|---|---|
| IGHV1-3 | 153 | -2.63 | -5.65 | 1.60E-08 | 9.36E-07 |
| IGLL1 | 306 | -2.06 | -5.65 | 1.59E-08 | 9.36E-07 |
| IGLJ3 | 102 | -2.58 | -5.63 | 1.80E-08 | 1.04E-06 |
| ITGAL | 1102 | -1.06 | -5.63 | 1.81E-08 | 1.05E-06 |
| IGKC | 23344 | -2.68 | -5.61 | 2.01E-08 | 1.15E-06 |
| GPR183 | 234 | -1.29 | -5.60 | 2.11E-08 | 1.20E-06 |
| IGKV2-28 | 772 | -2.72 | -5.59 | 2.26E-08 | 1.28E-06 |
| GGTA1 | 120 | -1.36 | -5.59 | 2.27E-08 | 1.28E-06 |
| IGKV1-39 | 1287 | -3.04 | -5.59 | 2.30E-08 | 1.30E-06 |
| IGKV2D-28 | 774 | -2.71 | -5.58 | 2.44E-08 | 1.37E-06 |
| PLPPR4 | 289 | -1.77 | -5.58 | 2.45E-08 | 1.37E-06 |
| IGKV1-12 | 672 | -2.91 | -5.56 | 2.63E-08 | 1.45E-06 |
| ITK | 315 | -1.41 | -5.56 | 2.63E-08 | 1.45E-06 |
| TNFSF13B | 182 | -1.52 | -5.56 | 2.68E-08 | 1.47E-06 |
| MXRA5 | 1567 | -1.59 | -5.56 | 2.72E-08 | 1.48E-06 |
| IGKV3-11 | 1105 | -2.67 | -5.53 | 3.26E-08 | 1.75E-06 |
| IGLV1-36 | 211 | -2.68 | -5.52 | 3.41E-08 | 1.83E-06 |
| FAM30A | 135 | -2.14 | -5.51 | 3.59E-08 | 1.92E-06 |
| IGKV1D-16 | 506 | -2.97 | -5.51 | 3.68E-08 | 1.97E-06 |
| NREP | 268 | -1.33 | -5.48 | 4.16E-08 | 2.19E-06 |
| IGHV1-24 | 187 | -2.82 | -5.47 | 4.53E-08 | 2.34E-06 |
| SLC8A1 | 473 | -1.28 | -5.46 | 4.74E-08 | 2.44E-06 |
| CD84 | 373 | -1.27 | -5.45 | 4.94E-08 | 2.54E-06 |
| CD48 | 435 | -1.40 | -5.45 | 5.14E-08 | 2.63E-06 |
| IGKV1D-12 | 661 | -2.87 | -5.44 | 5.27E-08 | 2.69E-06 |
| CD52 | 634 | -1.34 | -5.42 | 5.83E-08 | 2.94E-06 |
| IGKV2D-30 | 282 | -2.86 | -5.42 | 6.06E-08 | 3.03E-06 |
| MAP4K1 | 301 | -1.07 | -5.42 | 6.12E-08 | 3.05E-06 |

| | | | | | |
|---|---|---|---|---|---|
| GPR65 | 130 | -1.35 | -5.41 | 6.20E-08 | 3.09E-06 |
| IGKV3D-11 | 896 | -2.69 | -5.41 | 6.45E-08 | 3.19E-06 |
| SFRP4 | 611 | -3.05 | -5.40 | 6.58E-08 | 3.23E-06 |
| IGKV3D-20 | 798 | -2.78 | -5.40 | 6.63E-08 | 3.24E-06 |
| FGL2 | 1696 | -1.31 | -5.40 | 6.70E-08 | 3.26E-06 |
| ARHGAP15 | 232 | -1.28 | -5.39 | 7.18E-08 | 3.47E-06 |
| WDFY4 | 396 | -1.37 | -5.38 | 7.35E-08 | 3.54E-06 |
| TMC8 | 733 | -1.25 | -5.37 | 7.73E-08 | 3.71E-06 |
| IGHV3-23 | 906 | -2.77 | -5.34 | 9.44E-08 | 4.45E-06 |
| IGLV1-44 | 731 | -2.62 | -5.33 | 9.55E-08 | 4.48E-06 |
| FCRL5 | 100 | -2.24 | -5.33 | 9.78E-08 | 4.56E-06 |
| IGLV3-1 | 461 | -2.41 | -5.32 | 1.03E-07 | 4.76E-06 |
| LTB | 185 | -1.86 | -5.32 | 1.04E-07 | 4.83E-06 |
| IGHV3-15 | 305 | -2.71 | -5.30 | 1.14E-07 | 5.27E-06 |
| IGKV2-24 | 164 | -2.75 | -5.30 | 1.17E-07 | 5.36E-06 |
| ITIH5 | 850 | -1.89 | -5.29 | 1.20E-07 | 5.50E-06 |
| MUC13 | 2187 | -1.32 | -5.29 | 1.21E-07 | 5.52E-06 |
| IGKV3-20 | 1887 | -2.72 | -5.29 | 1.22E-07 | 5.55E-06 |
| TPSAB1 | 1466 | -1.48 | -5.28 | 1.27E-07 | 5.69E-06 |
| IGKV2-30 | 344 | -2.83 | -5.27 | 1.33E-07 | 5.96E-06 |
| MZB1 | 299 | -1.71 | -5.27 | 1.34E-07 | 5.98E-06 |
| DIO2 | 843 | -2.15 | -5.27 | 1.34E-07 | 5.99E-06 |
| IGHV3-74 | 299 | -2.53 | -5.24 | 1.63E-07 | 7.21E-06 |
| COL6A6 | 192 | -1.72 | -5.24 | 1.65E-07 | 7.23E-06 |
| IGHV4-61 | 624 | -2.44 | -5.23 | 1.70E-07 | 7.47E-06 |
| VASH1 | 655 | -1.08 | -5.21 | 1.87E-07 | 8.07E-06 |
| IGHV4-34 | 248 | -2.45 | -5.20 | 1.94E-07 | 8.31E-06 |
| NA | 289 | -1.36 | -5.20 | 2.00E-07 | 8.55E-06 |

| | | | | | |
|---|---|---|---|---|---|
| PDE4B | 265 | -1.30 | -5.19 | 2.11E-07 | 9.00E-06 |
| GZMH | 134 | -1.44 | -5.19 | 2.14E-07 | 9.07E-06 |
| IGHV3-49 | 224 | -2.73 | -5.18 | 2.18E-07 | 9.22E-06 |
| TPSB2 | 1643 | -1.47 | -5.18 | 2.21E-07 | 9.33E-06 |
| PARP15 | 244 | -1.11 | -5.18 | 2.22E-07 | 9.33E-06 |
| CD247 | 226 | -1.24 | -5.17 | 2.32E-07 | 9.61E-06 |
| IGHV4-39 | 523 | -2.51 | -5.17 | 2.36E-07 | 9.74E-06 |
| IGHV3-33 | 980 | -2.65 | -5.17 | 2.38E-07 | 9.81E-06 |
| DOCK2 | 679 | -1.24 | -5.16 | 2.45E-07 | 1.01E-05 |
| TBC1D10C | 441 | -1.09 | -5.16 | 2.50E-07 | 1.03E-05 |
| IGLV1-47 | 660 | -2.50 | -5.15 | 2.56E-07 | 1.05E-05 |
| ZBP1 | 101 | -1.51 | -5.15 | 2.60E-07 | 1.06E-05 |
| ROBO2 | 365 | -1.92 | -5.15 | 2.63E-07 | 1.07E-05 |
| GBP5 | 1043 | -1.17 | -5.14 | 2.71E-07 | 1.10E-05 |
| GZMA | 253 | -1.21 | -5.13 | 2.89E-07 | 1.17E-05 |
| IGHV5-51 | 316 | -2.56 | -5.13 | 2.93E-07 | 1.18E-05 |
| IL32 | 775 | -1.55 | -5.12 | 3.01E-07 | 1.21E-05 |
| IGLV2-18 | 357 | -2.53 | -5.11 | 3.23E-07 | 1.28E-05 |
| CTSW | 175 | -1.24 | -5.11 | 3.27E-07 | 1.29E-05 |
| ABI3BP | 1371 | -1.57 | -5.11 | 3.30E-07 | 1.30E-05 |
| PLEK | 620 | -1.18 | -5.10 | 3.45E-07 | 1.35E-05 |
| CXCL9 | 1772 | -2.90 | -5.09 | 3.54E-07 | 1.37E-05 |
| IRF4 | 217 | -1.78 | -5.08 | 3.85E-07 | 1.48E-05 |
| ADAM19 | 400 | -1.61 | -5.07 | 3.90E-07 | 1.49E-05 |
| IGHV3-20 | 192 | -2.67 | -5.07 | 3.94E-07 | 1.50E-05 |
| IGHV3-13 | 179 | -2.51 | -5.07 | 4.00E-07 | 1.52E-05 |
| FYB1 | 777 | -1.29 | -5.06 | 4.21E-07 | 1.59E-05 |
| MAF | 331 | -1.01 | -5.03 | 4.91E-07 | 1.82E-05 |

| | | | | | |
|---|---|---|---|---|---|
| IGKV1-33 | 744 | -2.82 | -5.02 | 5.11E-07 | 1.88E-05 |
| IGKV1D-33 | 746 | -2.81 | -5.01 | 5.53E-07 | 2.01E-05 |
| CD28 | 114 | -1.90 | -5.00 | 5.66E-07 | 2.06E-05 |
| IGKV1D-13 | 393 | -2.65 | -5.00 | 5.72E-07 | 2.08E-05 |
| CD22 | 154 | -1.64 | -4.98 | 6.39E-07 | 2.31E-05 |
| MCOLN2 | 210 | -1.08 | -4.97 | 6.73E-07 | 2.41E-05 |
| SEPTIN1 | 337 | -1.09 | -4.97 | 6.73E-07 | 2.41E-05 |
| TGFB3 | 627 | -1.00 | -4.97 | 6.80E-07 | 2.43E-05 |
| IGKV1-5 | 1078 | -2.60 | -4.97 | 6.83E-07 | 2.43E-05 |
| JAML | 795 | -1.13 | -4.96 | 6.91E-07 | 2.46E-05 |
| TNFSF14 | 189 | -1.11 | -4.96 | 7.06E-07 | 2.50E-05 |
| IGLV2-34 | 320 | -2.26 | -4.94 | 7.85E-07 | 2.75E-05 |
| IGHV3-7 | 662 | -2.70 | -4.92 | 8.44E-07 | 2.92E-05 |
| DCHS2 | 155 | -1.36 | -4.92 | 8.44E-07 | 2.92E-05 |
| LAIR1 | 382 | -1.07 | -4.92 | 8.67E-07 | 2.99E-05 |
| HLA-DOA | 1286 | -1.03 | -4.92 | 8.66E-07 | 2.99E-05 |
| IKZF1 | 697 | -1.10 | -4.92 | 8.86E-07 | 3.05E-05 |
| LCP1 | 3622 | -1.18 | -4.90 | 9.49E-07 | 3.23E-05 |
| UBD | 1644 | -1.66 | -4.90 | 9.71E-07 | 3.30E-05 |
| COL14A1 | 1515 | -1.60 | -4.88 | 1.03E-06 | 3.50E-05 |
| PARVG | 535 | -1.14 | -4.88 | 1.04E-06 | 3.52E-05 |
| HLA-DOB | 121 | -1.40 | -4.86 | 1.20E-06 | 3.97E-05 |
| MS4A6A | 704 | -1.07 | -4.85 | 1.22E-06 | 4.01E-05 |
| IGHV1-2 | 339 | -2.66 | -4.85 | 1.21E-06 | 4.01E-05 |
| CARMIL2 | 163 | -1.29 | -4.84 | 1.33E-06 | 4.35E-05 |
| HSPA7 | 295 | -1.18 | -4.83 | 1.34E-06 | 4.37E-05 |
| OASL | 178 | -1.40 | -4.83 | 1.37E-06 | 4.46E-05 |
| IGHV4-28 | 141 | -2.63 | -4.83 | 1.37E-06 | 4.48E-05 |

| | | | | | |
|---|---|---|---|---|---|
| TRAF3IP3 | 415 | -1.08 | -4.82 | 1.42E-06 | 4.60E-05 |
| GNG2 | 328 | -1.20 | -4.82 | 1.46E-06 | 4.71E-05 |
| IGHV4-59 | 655 | -2.38 | -4.81 | 1.53E-06 | 4.89E-05 |
| DOK2 | 182 | -1.12 | -4.80 | 1.57E-06 | 5.02E-05 |
| IGHV3-30 | 938 | -2.53 | -4.77 | 1.80E-06 | 5.66E-05 |
| IGLV2-23 | 1054 | -2.19 | -4.75 | 2.05E-06 | 6.32E-05 |
| HMCN1 | 1660 | -1.38 | -4.73 | 2.28E-06 | 6.97E-05 |
| AGAP2 | 141 | -1.19 | -4.73 | 2.28E-06 | 6.97E-05 |
| P2RY13 | 209 | -1.33 | -4.70 | 2.54E-06 | 7.63E-05 |
| IGLC7 | 666 | -2.35 | -4.69 | 2.68E-06 | 8.02E-05 |
| IGHV2-5 | 149 | -2.67 | -4.68 | 2.83E-06 | 8.42E-05 |
| IFI44L | 1914 | -1.28 | -4.67 | 2.95E-06 | 8.73E-05 |
| TPSD1 | 222 | -1.39 | -4.67 | 3.00E-06 | 8.84E-05 |
| MMP9 | 101 | -2.15 | -4.67 | 3.02E-06 | 8.87E-05 |
| IGLC3 | 8964 | -2.26 | -4.66 | 3.17E-06 | 9.24E-05 |
| IGLV3-9 | 121 | -2.63 | -4.65 | 3.25E-06 | 9.41E-05 |
| CORO1A | 1616 | -1.12 | -4.64 | 3.48E-06 | 9.97E-05 |
| CD7 | 197 | -1.33 | -4.62 | 3.75E-06 | 0.0001 |
| CCL19 | 838 | -2.77 | -4.58 | 4.59E-06 | 0.0001 |
| FGF1 | 139 | -1.50 | -4.58 | 4.69E-06 | 0.0001 |
| LAMA4 | 1644 | -1.26 | -4.57 | 4.78E-06 | 0.0001 |
| COL10A1 | 203 | -1.26 | -4.57 | 4.91E-06 | 0.0001 |
| EXOC3L4 | 157 | -1.02 | -4.57 | 4.99E-06 | 0.0001 |
| OLFML2B | 324 | -1.19 | -4.52 | 6.05E-06 | 0.0002 |
| IGLV2-11 | 811 | -2.36 | -4.52 | 6.06E-06 | 0.0002 |
| GBP4 | 2156 | -1.01 | -4.51 | 6.58E-06 | 0.0002 |
| SLITRK5 | 116 | -1.19 | -4.50 | 6.83E-06 | 0.0002 |
| RAC2 | 950 | -1.06 | -4.49 | 7.00E-06 | 0.0002 |

| | | | | | |
|---|---|---|---|---|---|
| TFEC | 124 | -1.12 | -4.45 | 8.76E-06 | 0.0002 |
| IGLV7-46 | 101 | -2.68 | -4.44 | 9.04E-06 | 0.0002 |
| PSTPIP1 | 195 | -1.10 | -4.43 | 9.36E-06 | 0.0002 |
| IGLV2-14 | 1781 | -2.27 | -4.43 | 9.49E-06 | 0.0002 |
| IGLL5 | 1198 | -2.26 | -4.41 | 1.03E-05 | 0.0003 |
| IGHA2 | 7043 | -2.08 | -4.40 | 1.10E-05 | 0.0003 |
| BTK | 207 | -1.15 | -4.40 | 1.10E-05 | 0.0003 |
| IGHA1 | 21057 | -1.97 | -4.39 | 1.13E-05 | 0.0003 |
| COL1A2 | 4278 | -2.00 | -4.37 | 1.24E-05 | 0.0003 |
| MS4A1 | 361 | -1.48 | -4.36 | 1.30E-05 | 0.0003 |
| IGLV2-8 | 858 | -2.32 | -4.35 | 1.34E-05 | 0.0003 |
| IGLV6-57 | 182 | -2.41 | -4.34 | 1.41E-05 | 0.0003 |
| VCAM1 | 351 | -2.08 | -4.34 | 1.42E-05 | 0.0003 |
| GUCY1A1 | 779 | -1.28 | -4.34 | 1.43E-05 | 0.0003 |
| ECM2 | 493 | -1.11 | -4.31 | 1.60E-05 | 0.0004 |
| MRC2 | 872 | -1.26 | -4.31 | 1.67E-05 | 0.0004 |
| AIF1 | 471 | -1.09 | -4.30 | 1.71E-05 | 0.0004 |
| GUCY1A2 | 178 | -1.80 | -4.27 | 1.98E-05 | 0.0004 |
| IGLV3-19 | 292 | -2.58 | -4.26 | 2.03E-05 | 0.0005 |
| IGLC2 | 11569 | -2.16 | -4.25 | 2.15E-05 | 0.0005 |
| LINC00861 | 116 | -1.33 | -4.25 | 2.17E-05 | 0.0005 |
| ITLN1 | 171 | -1.70 | -4.24 | 2.28E-05 | 0.0005 |
| CYBB | 986 | -1.16 | -4.23 | 2.35E-05 | 0.0005 |
| MYO1G | 305 | -1.08 | -4.23 | 2.36E-05 | 0.0005 |
| HCST | 131 | -1.08 | -4.22 | 2.42E-05 | 0.0005 |
| IGLV3-25 | 461 | -2.37 | -4.21 | 2.59E-05 | 0.0006 |
| CXCL14 | 1612 | -1.69 | -4.20 | 2.67E-05 | 0.0006 |
| ST8SIA6 | 120 | -1.01 | -4.20 | 2.72E-05 | 0.0006 |

| | | | | | |
|---|---|---|---|---|---|
| CALHM6 | 166 | -1.11 | -4.19 | 2.85E-05 | 0.0006 |
| TNFAIP8L2 | 112 | -1.00 | -4.18 | 2.93E-05 | 0.0006 |
| SLCO2B1 | 822 | -1.02 | -4.17 | 2.99E-05 | 0.0006 |
| GAPT | 104 | -1.08 | -4.17 | 3.00E-05 | 0.0006 |
| IGLV3-21 | 537 | -2.08 | -4.15 | 3.35E-05 | 0.0007 |
| POSTN | 3332 | -1.07 | -4.14 | 3.45E-05 | 0.0007 |
| COL3A1 | 4570 | -2.19 | -4.14 | 3.50E-05 | 0.0007 |
| LUM | 1511 | -2.45 | -4.13 | 3.56E-05 | 0.0007 |
| COL1A1 | 2141 | -1.21 | -4.13 | 3.62E-05 | 0.0007 |
| LILRB1 | 247 | -1.17 | -4.13 | 3.65E-05 | 0.0007 |
| FREM1 | 220 | -1.59 | -4.07 | 4.66E-05 | 0.0009 |
| PTGDS | 1569 | -2.07 | -4.06 | 4.83E-05 | 0.001 |
| CXCL10 | 1155 | -2.14 | -4.06 | 4.92E-05 | 0.001 |
| NTRK2 | 298 | -1.13 | -3.99 | 6.56E-05 | 0.001 |
| CCL21 | 326 | -2.04 | -3.98 | 7.03E-05 | 0.001 |
| SIGLEC1 | 217 | -1.11 | -3.97 | 7.21E-05 | 0.001 |
| P2RX7 | 112 | -1.00 | -3.96 | 7.39E-05 | 0.001 |
| EDNRA | 635 | -1.04 | -3.91 | 9.37E-05 | 0.002 |
| HGF | 151 | -1.61 | -3.90 | 9.68E-05 | 0.002 |
| IGHD | 1230 | -1.83 | -3.87 | 0.0001 | 0.002 |
| ENPP2 | 666 | -1.24 | -3.85 | 0.0001 | 0.002 |
| F2RL2 | 166 | -1.28 | -3.79 | 0.0001 | 0.002 |
| SLC2A14 | 123 | -1.13 | -3.79 | 0.0002 | 0.002 |
| NKG7 | 230 | -1.22 | -3.78 | 0.0002 | 0.003 |
| CXCL12 | 1272 | -1.58 | -3.77 | 0.0002 | 0.003 |
| PTGS1 | 437 | -1.05 | -3.77 | 0.0002 | 0.003 |
| CTHRC1 | 101 | -1.38 | -3.73 | 0.0002 | 0.003 |
| CST7 | 123 | -1.09 | -3.69 | 0.0002 | 0.003 |

| | | | | | |
|---|---|---|---|---|---|
| PRRX1 | 757 | -1.26 | -3.66 | 0.0003 | 0.004 |
| MATN2 | 1086 | -1.02 | -3.66 | 0.0003 | 0.004 |
| F13A1 | 1017 | -1.65 | -3.66 | 0.0003 | 0.004 |
| CD79A | 220 | -1.56 | -3.64 | 0.0003 | 0.004 |
| ASPN | 719 | -1.63 | -3.62 | 0.0003 | 0.004 |
| LSAMP | 674 | -1.32 | -3.61 | 0.0003 | 0.004 |
| LST1 | 257 | -1.01 | -3.54 | 0.0004 | 0.005 |
| TMEM119 | 351 | -1.52 | -3.51 | 0.0004 | 0.006 |
| TFF1 | 113 | -1.30 | -3.51 | 0.0004 | 0.006 |
| CSF2RB | 701 | -1.18 | -3.51 | 0.0005 | 0.006 |
| COL5A1 | 923 | -1.42 | -3.50 | 0.0005 | 0.006 |
| OLFML1 | 306 | -1.13 | -3.50 | 0.0005 | 0.006 |
| CHI3L2 | 187 | -1.52 | -3.50 | 0.0005 | 0.006 |
| LDB2 | 637 | -1.11 | -3.45 | 0.0006 | 0.007 |
| JAK3 | 641 | -1.01 | -3.45 | 0.0006 | 0.007 |
| SGIP1 | 107 | -1.50 | -3.44 | 0.0006 | 0.007 |
| CXCL11 | 313 | -2.05 | -3.43 | 0.0006 | 0.008 |
| CPNE5 | 140 | -1.01 | -3.41 | 0.0007 | 0.008 |
| ADGRA2 | 810 | -1.35 | -3.40 | 0.0007 | 0.008 |
| FNDC1 | 113 | -1.39 | -3.39 | 0.0007 | 0.008 |
| IGHM | 2170 | -1.54 | -3.38 | 0.0007 | 0.009 |
| EGFLAM | 116 | -1.27 | -3.36 | 0.0008 | 0.009 |
| PLA2G4C | 205 | -1.09 | -3.35 | 0.0008 | 0.010 |
| CEACAM5 | 2470 | -1.07 | -3.33 | 0.0009 | 0.010 |
| JAM2 | 720 | -1.10 | -3.28 | 0.0010 | 0.012 |
| CLIC2 | 381 | -1.00 | -3.27 | 0.0011 | 0.012 |
| PI16 | 645 | -1.54 | -3.23 | 0.0012 | 0.013 |
| FAM111B | 100 | -1.16 | -3.21 | 0.0013 | 0.014 |

| | | | | | |
|---|---|---|---|---|---|
| ABCB1 | 521 | -1.08 | -3.19 | 0.0014 | 0.015 |
| MMP10 | 3182 | -1.01 | -3.19 | 0.0014 | 0.015 |
| DPT | 1681 | -1.72 | -3.18 | 0.0014 | 0.015 |
| TSPAN18 | 337 | -1.04 | -3.18 | 0.0015 | 0.015 |
| COL15A1 | 2580 | -1.70 | -3.17 | 0.0015 | 0.016 |
| OGN | 554 | -1.47 | -3.15 | 0.0016 | 0.016 |
| EPHA3 | 338 | -1.13 | -3.15 | 0.0016 | 0.016 |
| FRZB | 237 | -1.27 | -3.12 | 0.0018 | 0.018 |
| CD248 | 132 | -1.18 | -3.08 | 0.0020 | 0.019 |
| GLI1 | 116 | -1.10 | -3.06 | 0.0022 | 0.021 |
| COL8A1 | 243 | -1.20 | -3.05 | 0.0023 | 0.021 |
| PREX2 | 323 | -1.35 | -3.01 | 0.0026 | 0.024 |
| CACNA1C | 281 | -1.04 | -2.99 | 0.0028 | 0.025 |
| BGN | 2401 | -1.49 | -2.98 | 0.0028 | 0.025 |
| COLEC12 | 198 | -1.02 | -2.98 | 0.0029 | 0.025 |
| SPON1 | 1268 | -1.14 | -2.98 | 0.0029 | 0.025 |
| CLEC3B | 216 | -1.17 | -2.98 | 0.0029 | 0.025 |
| SELE | 306 | -1.67 | -2.97 | 0.0029 | 0.026 |
| TEK | 353 | -1.15 | -2.92 | 0.0035 | 0.029 |
| FAT4 | 1574 | -1.15 | -2.86 | 0.0042 | 0.034 |
| FLRT2 | 373 | -1.46 | -2.85 | 0.0043 | 0.035 |
| IDO1 | 1607 | -1.46 | -2.85 | 0.0044 | 0.035 |
| HAS2 | 115 | -1.18 | -2.83 | 0.0047 | 0.037 |
| BCHE | 209 | -1.68 | -2.82 | 0.0048 | 0.038 |
| MEOX1 | 415 | -1.45 | -2.81 | 0.0049 | 0.038 |
| DNM3OS | 178 | -1.04 | -2.81 | 0.0049 | 0.038 |
| ZNF385D | 246 | -1.30 | -2.81 | 0.0050 | 0.038 |
| COL6A2 | 3477 | -1.39 | -2.79 | 0.0053 | 0.040 |

| | | | | | |
|---|---|---|---|---|---|
| SELP | 623 | -1.22 | -2.76 | 0.0057 | 0.043 |
| GREM2 | 293 | -1.12 | -2.76 | 0.0058 | 0.043 |
| HPSE2 | 200 | -1.43 | -2.74 | 0.0062 | 0.045 |
| MOXD1 | 438 | -1.21 | -2.73 | 0.0064 | 0.046 |

**Supplementary Table E5 Geneset enrichment analysis for top 10 differentially enriched gene ontology pathways in bronchial brushes**

Gene ontology (GO) pathways significantly differentially expressed after 4 weeks of inhaled fluticasone (treatment group only) using Limma. Pathways shown are censored at Benajmini Hochberg adjusted P < 0.05 and ordered by P value. Under figure *x* axis represents approximate gene ranks. NES, normalise enrichment score.

| Brushings | GO Pathway | Gene ranks | NES | p | p adj |
|---|---|---|---|---|---|
| Upregulated by ICS | Positive regulation of ERBB signaling pathway | | 1.93 | 0.0004276 | 0.0450544 |
| | Cytokine activity | | 1.59 | 0.00064 | 0.053437 |
| | Positive regulation of T cell mediated cytotoxicity | | 2.02 | 0.0008298 | 0.0626609 |
| | Regulation of B cell receptor signaling pathway | | 1.96 | 0.0019826 | 0.1082716 |
| | Cytokine mediated signaling pathway | | 1.39 | 0.0021439 | 0.1101276 |
| | Positive regulation of adaptive immune response | | 1.60 | 0.0028807 | 0.1314115 |
| | Regulation of T cell mediated cytotoxicity | | 1.86 | 0.0031884 | 0.1383193 |
| | Tumor necrosis factor mediated signaling pathway | | 1.56 | 0.0043835 | 0.1731496 |
| | Regulation of interferon gamma production | | 1.56 | 0.0047389 | 0.1766441 |
| | Mannosidase activity | | 1.87 | 0.0048731 | 0.1782097 |
| Downregulated by ICS | Adaptive immune response | | -1.99 | 4.291E-09 | 1.502E-05 |
| | Lymphocyte activation | | -1.91 | 6.788E-09 | 1.502E-05 |
| | Regulation of immune effector process | | -1.81 | 1.274E-08 | 1.879E-05 |
| | Immune effector process | | -1.81 | 2.633E-08 | 2.913E-05 |
| | Leukocyte activation | | -1.78 | 1.474E-07 | 0.0001304 |
| | Lymphocyte mediated immunity | | -2.04 | 1.793E-07 | 0.0001322 |
| | Leukocyte mediated immunity | | -1.94 | 4.771E-07 | 0.0003016 |
| | Sodium ion homoestasis | | -2.29 | 1.517E-06 | 0.0008388 |
| | Activation of immune response | | -1.67 | 2.8E-06 | 0.0013549 |
| | Positive regulation of cell activation | | -1.74 | 3.062E-06 | 0.0013549 |

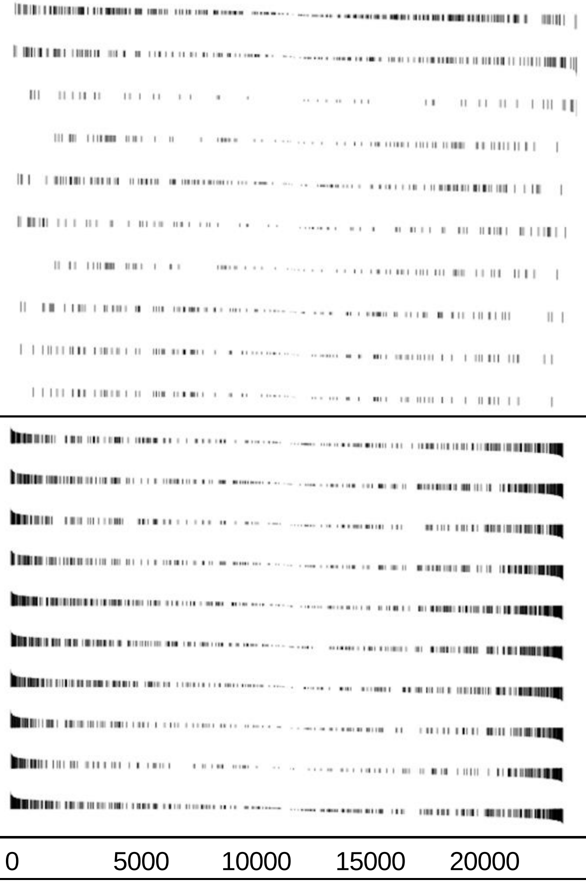

**Supplementary Table E6. Geneset enrichment analysis for top 10 differentially enriched gene ontology pathways in bronchial biopises.**

Gene ontology (GO) pathways significantly differentially expressed after 4 weeks of inhaled fluticasone (treatment group only) using Limma. Pathways shown are censored at Benajmini Hochberg adjusted $P < 0.05$ and ordered by P value. Under figure *x* axis represents approximate gene ranks. NES, normalise enrichment score.

| Biopsies | GO Pathway | Gene ranks | NES | p | p adj |
|---|---|---|---|---|---|
| Upregulated by ICS | Mitochondrial matrix | | 2.03 | 1.90711E-12 | 6.71695E-11 |
| | Oxidoreductase activity acting on the CH CH group of donors | | 2.50 | 5.46935E-09 | 1.1432E-07 |
| | Cofactor binding | | 1.98 | 1.12177E-08 | 2.1816E-07 |
| | Mitochondrial translation | | 2.31 | 1.53754E-08 | 2.88968E-07 |
| | Cilium organization | | 2.08 | 2.39225E-08 | 4.2974E-07 |
| | Organellar ribosome | | 2.31 | 3.52179E-08 | 6.03562E-07 |
| | DNA Packaging complex | | 2.25 | 6.07677E-08 | 9.95656E-07 |
| | Cellular respiration | | 2.12 | 1.22605E-07 | 1.83199E-06 |
| | Translational termination | | 2.25 | 1.6989E-07 | 2.41244E-06 |
| | Mitochondrial protein complex | | 2.12 | 3.30354E-07 | 4.42417E-06 |
| Downregulated by ICS | Regulation of cell activation | | -3.28 | 3.83652E-67 | 1.71607E-63 |
| | Adaptive immune response | | -3.45 | 3.00654E-57 | 5.21315E-54 |
| | Positive regulation of cell activation | | -3.38 | 3.49641E-57 | 5.21315E-54 |
| | Side of membrane | | -3.16 | 1.28859E-54 | 1.44097E-51 |
| | Activation of immune response | | -3.14 | 2.03234E-54 | 1.81814E-51 |
| | Immune effector process | | -3.07 | 3.35375E-53 | 2.50022E-50 |
| | Regulation of homotypic cell cell adhesion | | -3.24 | 2.82053E-50 | 1.80232E-47 |
| | Lymphocyte activation | | -3.19 | 9.17767E-49 | 5.13147E-46 |
| | Leukocyte activation | | -3.09 | 1.07142E-48 | 5.32495E-46 |
| | Regulation of cell cell adhesion | | -3.12 | 2.97918E-47 | 1.33259E-44 |

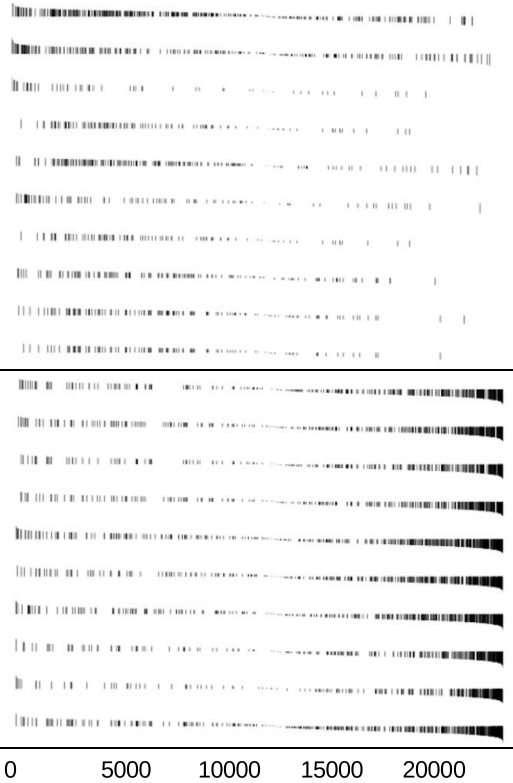

### Supplementary Figure E1

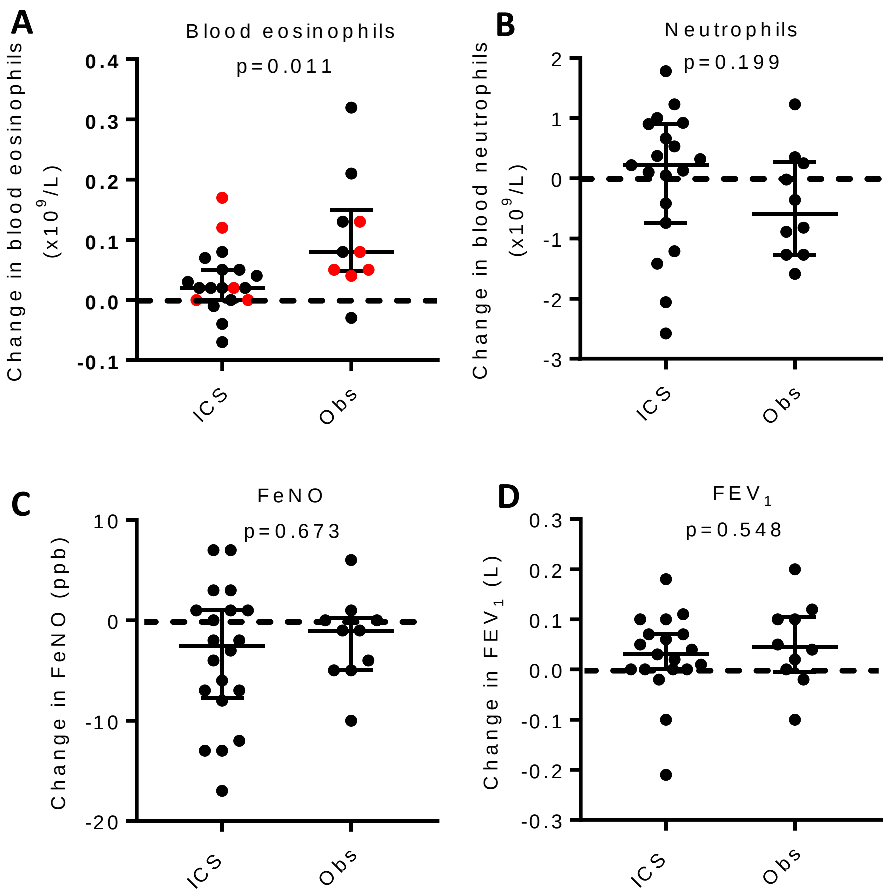


**Supplementary Figure E1**

The change in peripheral blood biomarkers and airway physiology on participants with available paired data before and after 4 weeks' treatment with inhaled fluticasone or without treatment. Changes in numbers of **A**) eosinophils, with atopic participants shown in red, or **B**) neutrophils in peripheral blood. Changes in **C**) fractional exhaled nitric oxide (FeNO) or **D**) forced expiratory volume in 1 second ($FEV_1$). Horizontal bars represent median (IQR), analysed by Mann Whitney U test.

### Supplementary Figure E2

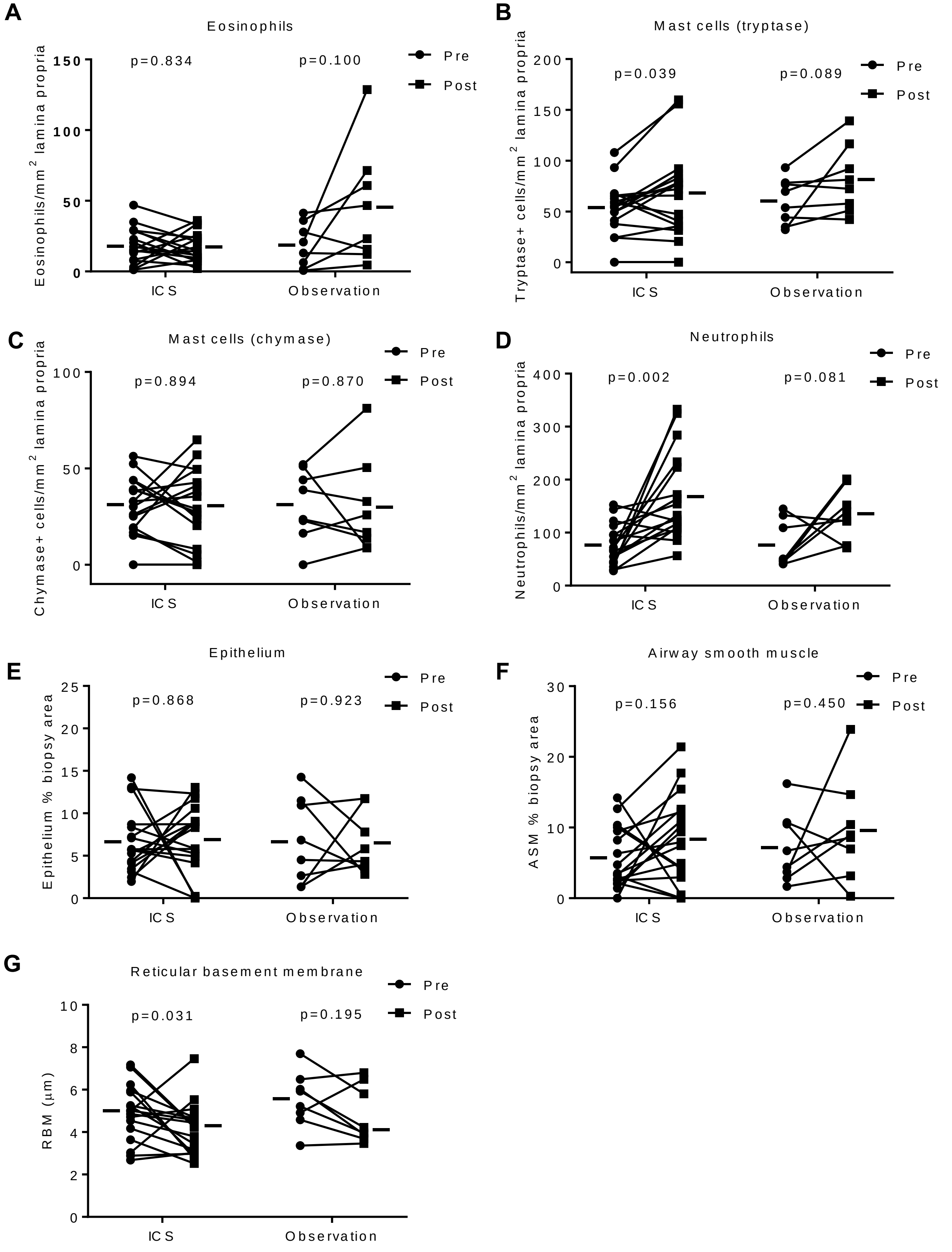

**Supplementary Figure E2 (above)**

Immunohistochemical analysis of the lamina propria showing cell counts and remodelling features from participants with available paired data before and after 4 weeks treatment with inhaled fluticasone or without treatment. Figure shows before and after data for numbers of **A**) eosinophils, with atopic participants shown in red, **B**) tryptase-positive mast cells, **C**) chymase-positive mast cells, or **D**) neutrophils, expressed in absolute counts/mm$^2$. Areas of **E**) epithelium or **F**) airway smooth muscle (ASM), expressed as a percentage of biopsy area. **G**) Measurements of reticular basement membrane (RBM) thickness. Horizontal bars represent mean (SD)(eosinophils, mast cells, neutrophils, ASM, epithelium) or median (IQR) (RBM), analysed by paired t test or Wilcoxon ranked pairs tests respectively.

### Supplementary Figure E3

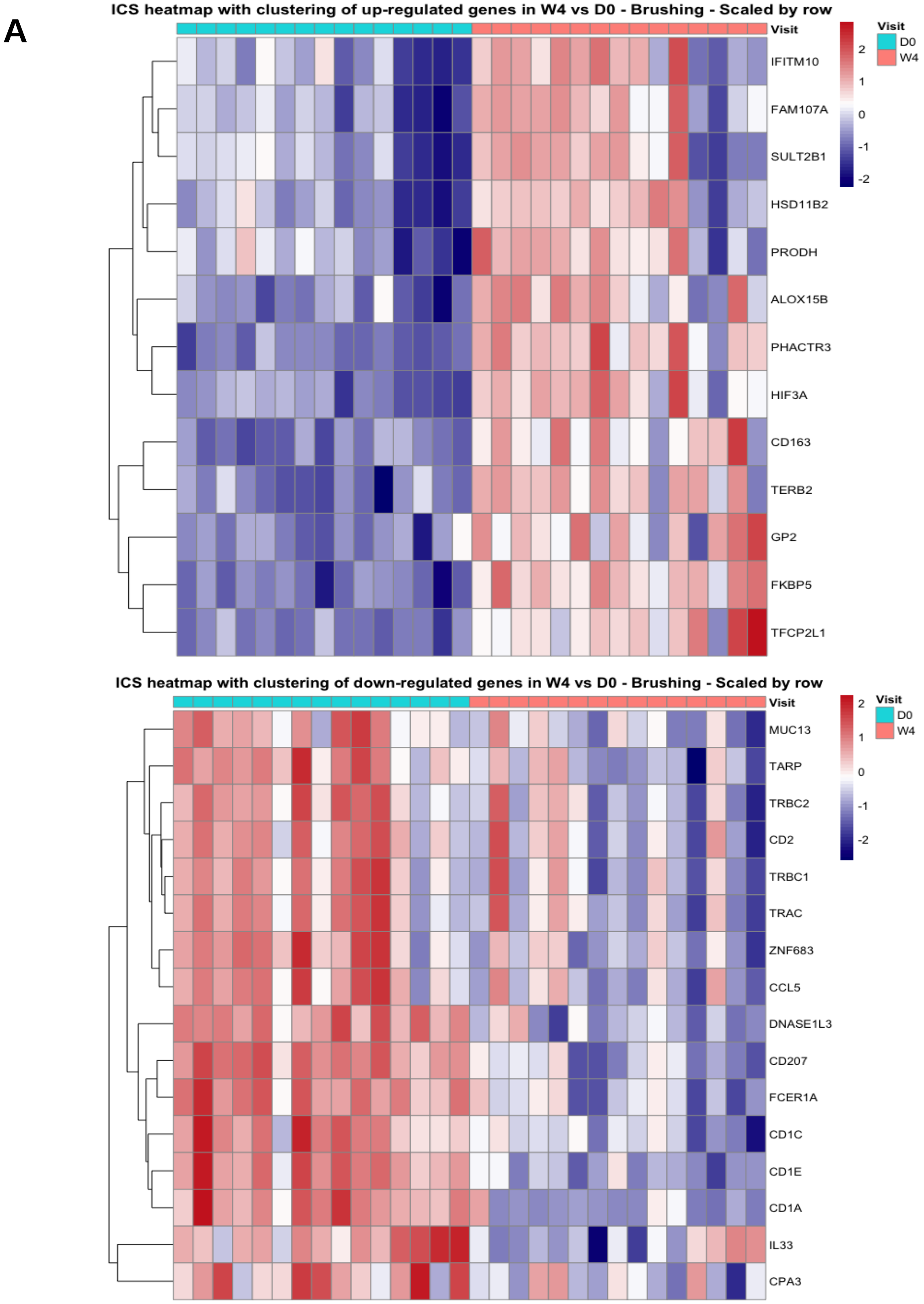


**Supplementary Figure E3.** Changes in gene expression measured by RNAseq in response to 4 weeks treatment with inhaled fluticasone. **A**) Bronchial brush heatmap representing the most differentially expressed genes (adjusted P<0.001, log2 fold change>0.5). **B**) Bronchial biopsy heatmap representing the most differentially expressed genes (adjusted P<0.001, log2 fold change>0.5).

### Supplementary Figure E3 continued

B

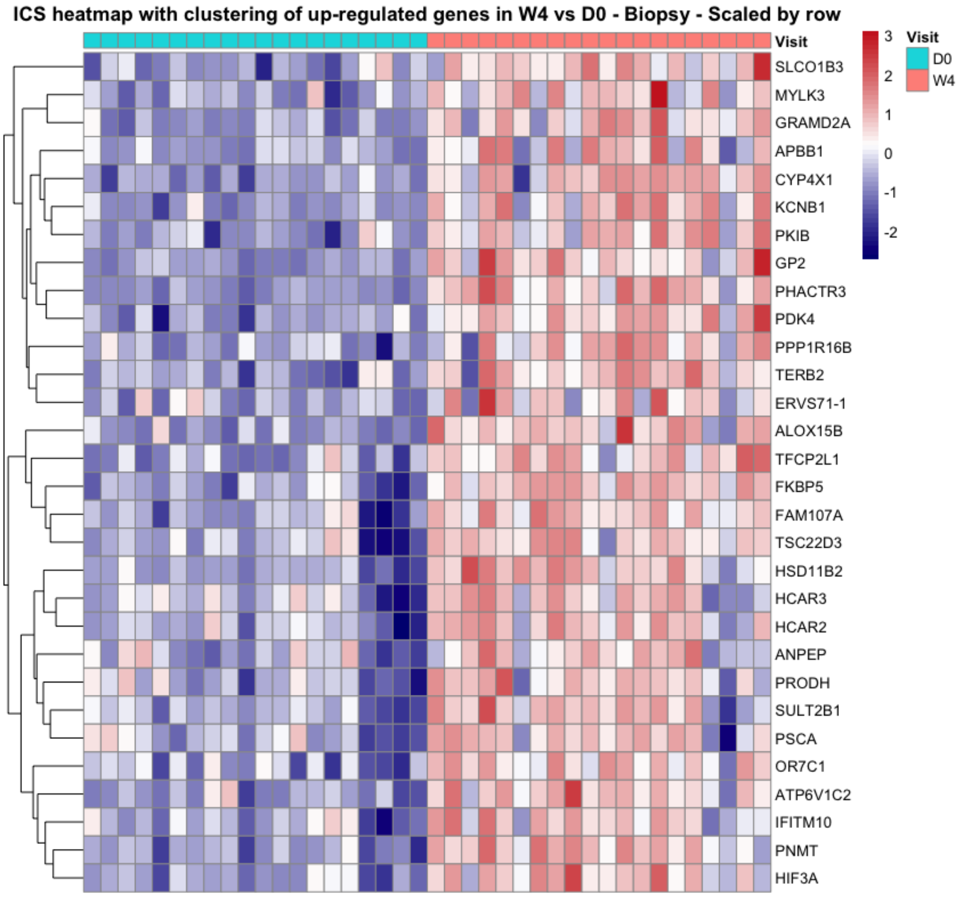


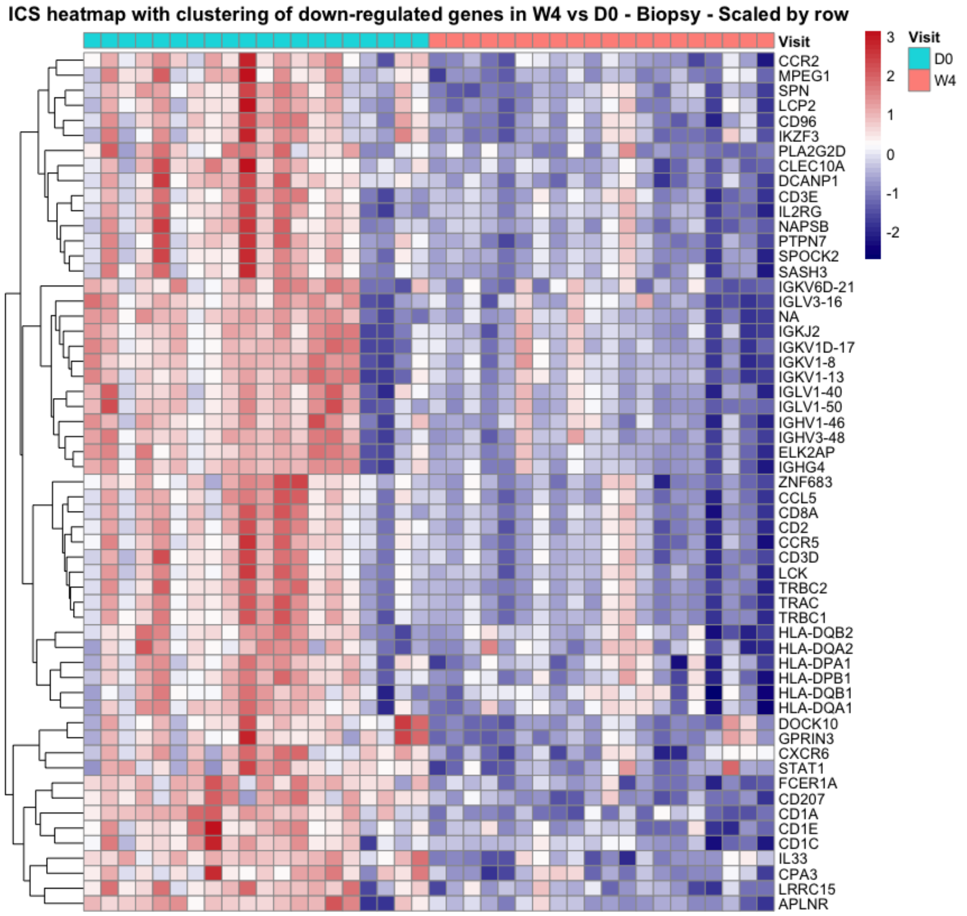

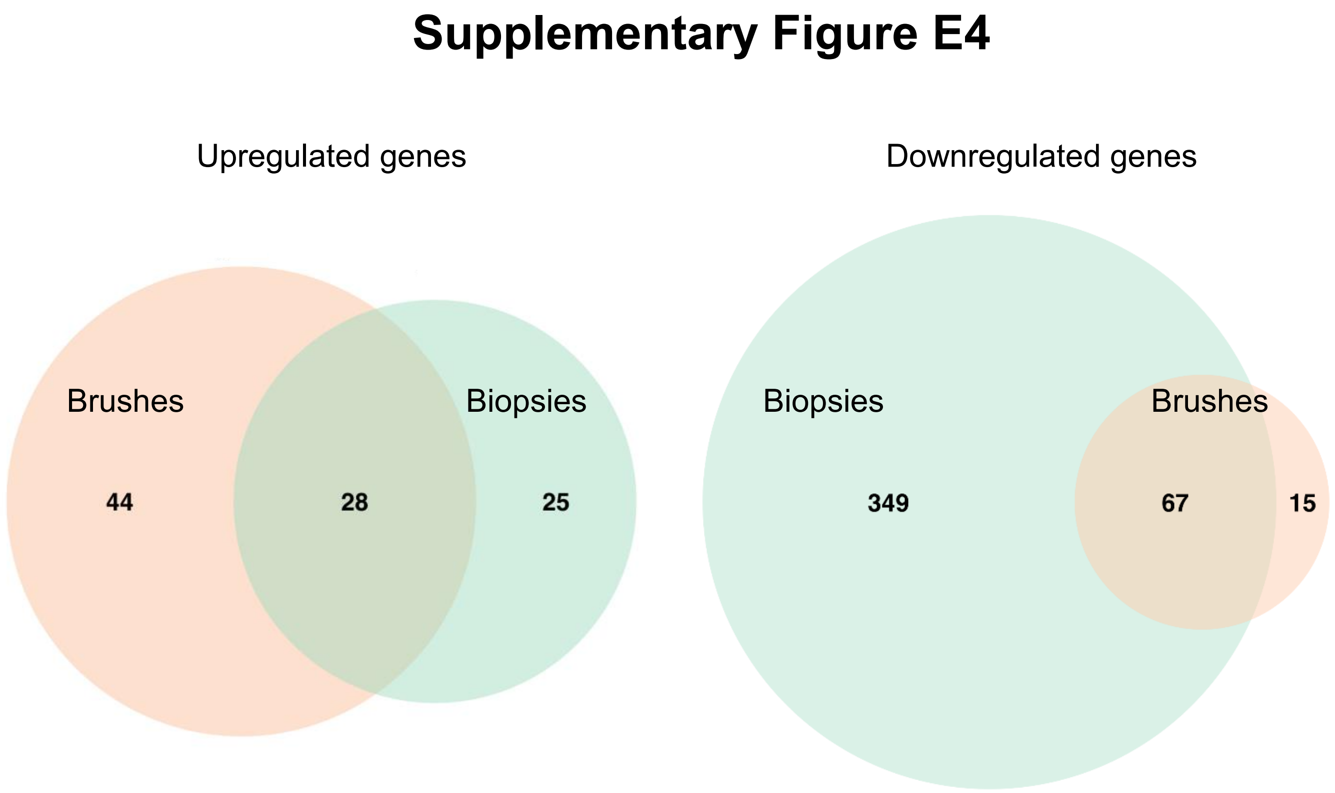


**Supplementary Figure E4**

Venn diagram of numbers of differentially expressed genes at week 4 versus baseline amongst those receiving inhaled fluticasone, showing genes **A**) upregulated and **B**) downregulated. Numbers generated with DeSeq2, with a significance threshold of log2 fold change > 1 and adjusted $p < 0.05$.

### Supplementary Figure E5

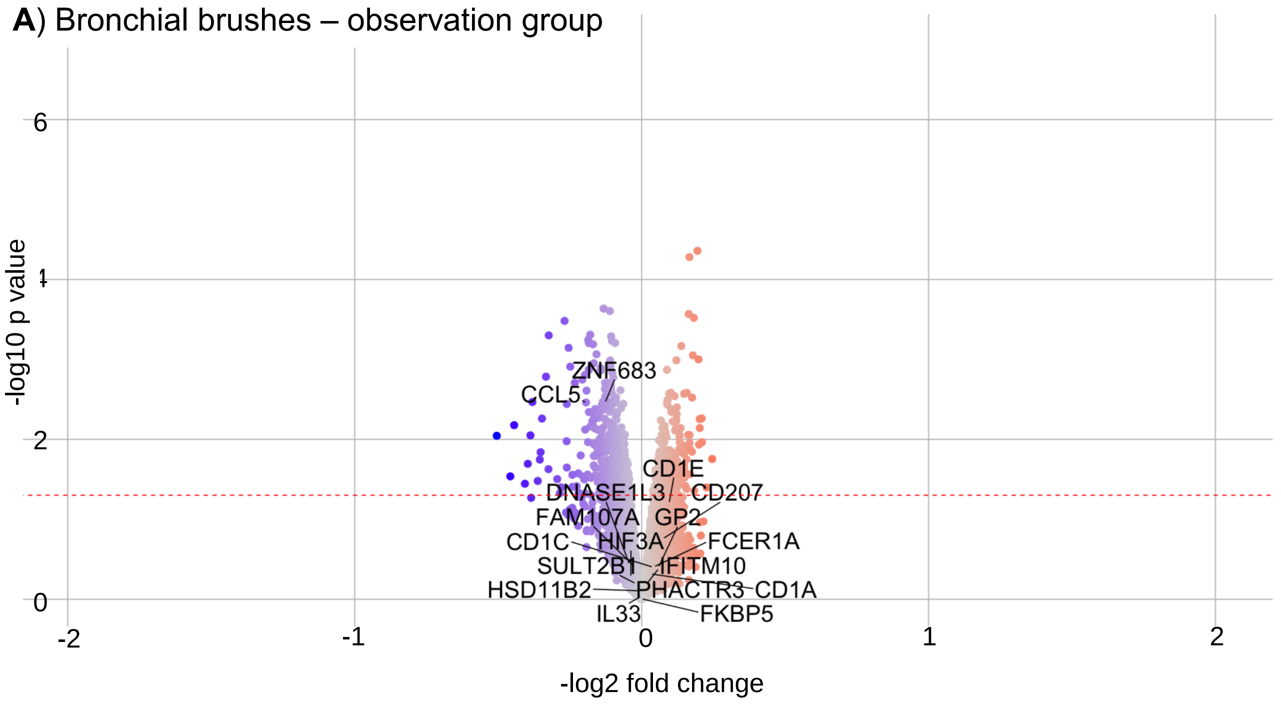


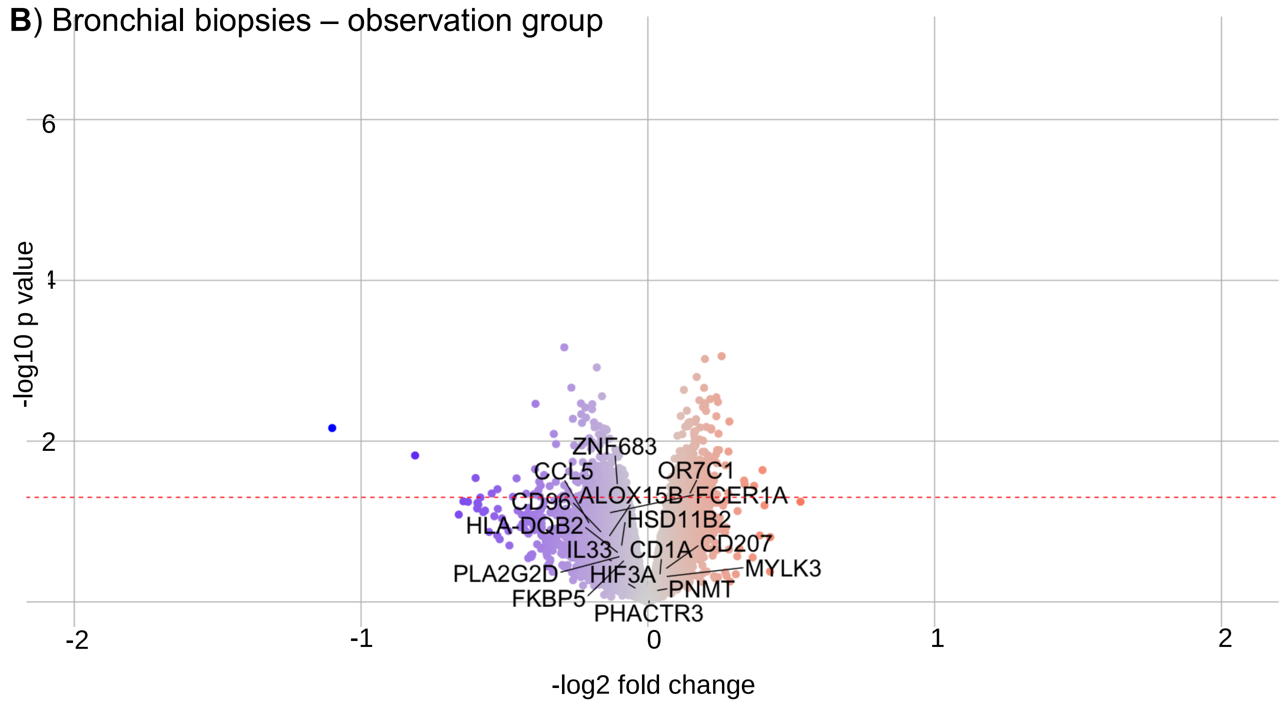


**Supplementary Figure E5**

Changes in gene expression measured by RNAseq following 4 weeks observation (control group). **A**) Bronchial brush volcano plot. **B**) Bronchial biopsy volcano plot. The labelled genes are a subset of those that were significantly changed by ICS. p values have not been adjusted.

### Supplementary Figure E6

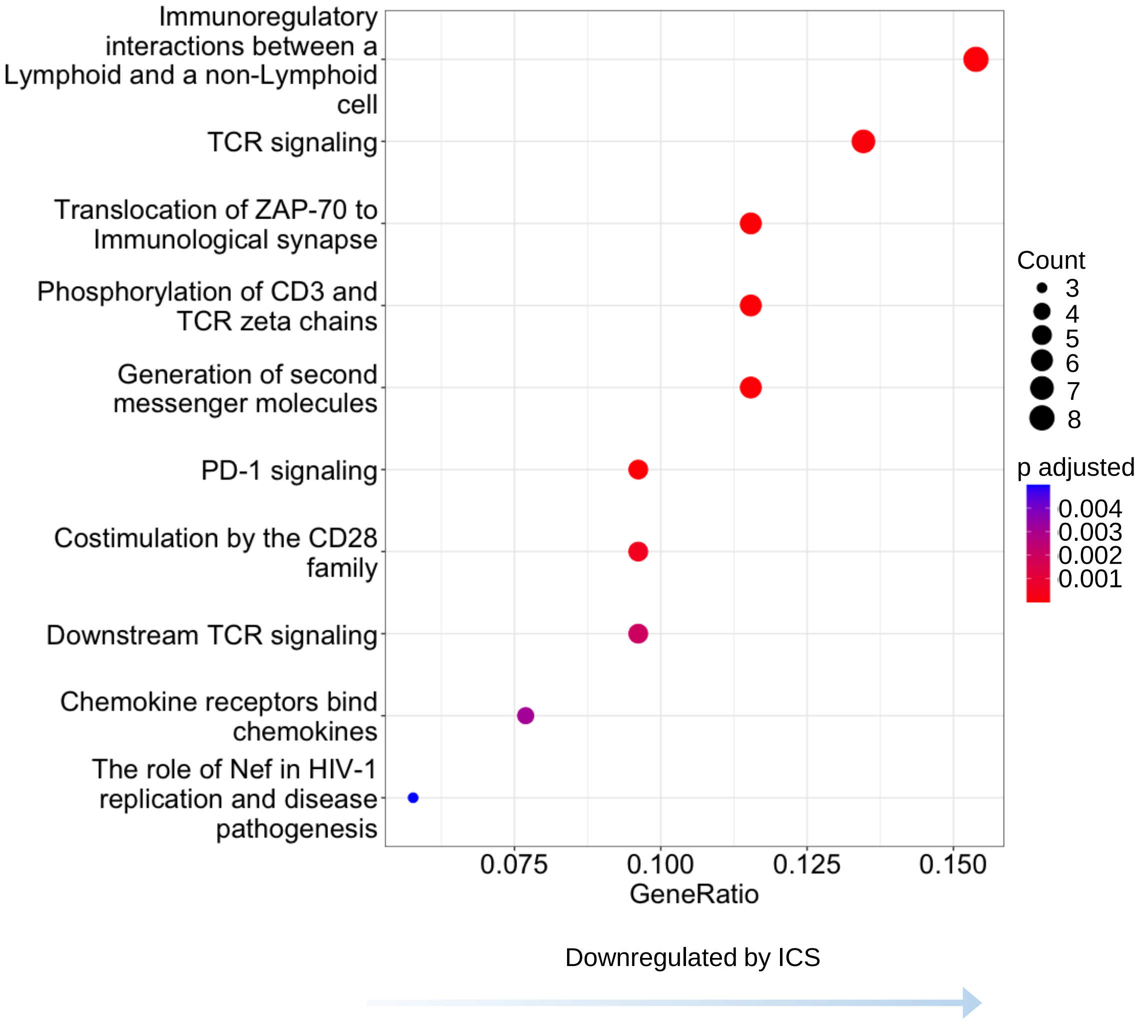


**Supplementary Figure E6. Reactome pathway analysis of bronchial brushings**

Reactome pathways significantly differentially enriched in bronchial brushing downregulated genes (log2 fold change >0, adjusted P value <0.25) after 4 weeks of inhaled fluticasone (treatment group only). x axis represents the gene ratio, the percentage of total differentially expressed genes in the given pathway; node size is proportional to number of genes differentially expressed in the pathway; node colour is proportional to Benjamini-Hochberg adjusted p value.

#### Supplementary Figure E7

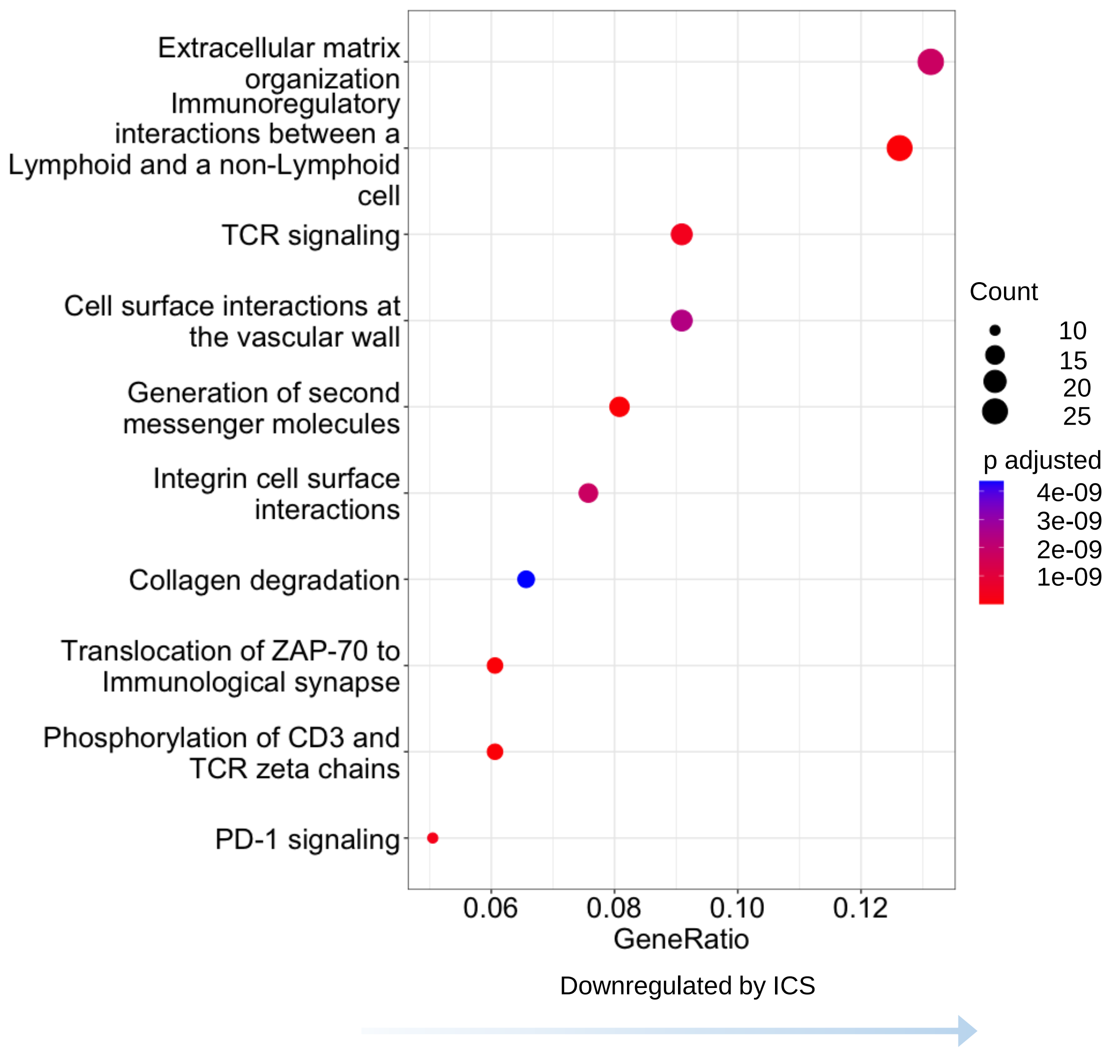


**Supplementary Figure E7. Reactome pathway analysis of bronchial biopsies**

Reactome pathways significantly differentially enriched in bronchial biopsy downregulated genes (log2 fold change >0, adjusted P value <0.25) after 4 weeks of inhaled fluticasone (treatment group only). x axis represents the gene ratio, the percentage of total differentially expressed genes in the given pathway; node size is proportional to number of genes differentially expressed in the pathway; node colour is proportional to Benjamini-Hochberg adjusted p value.

### Supplementary Figure E8

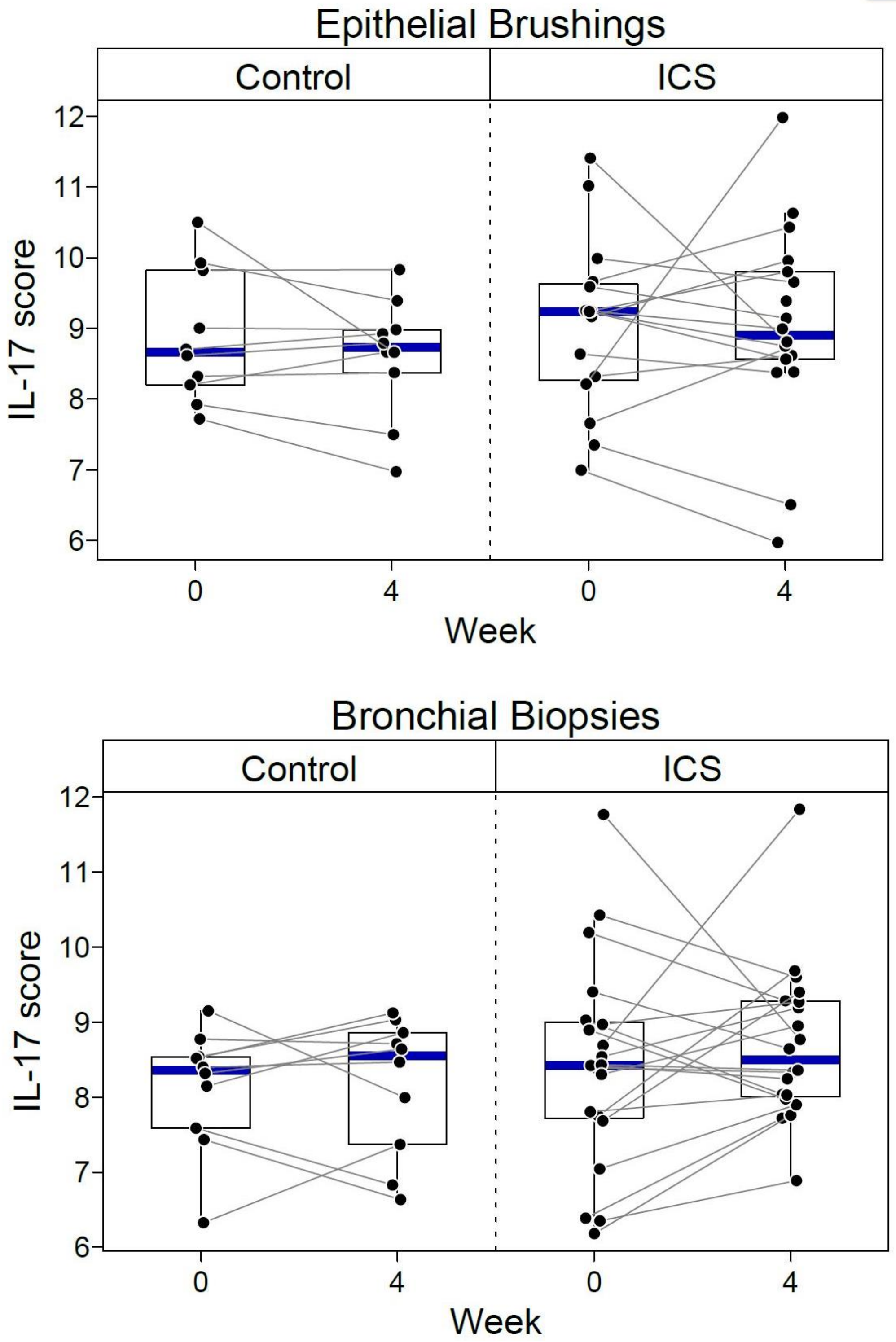


**Supplementary Figure E8.** Expression of an IL-17-dependent gene signature in bronchial brushes and biopsies in response to 4 weeks treatment with inhaled fluticasone, or observation (control).

#### Supplementary Figure E9

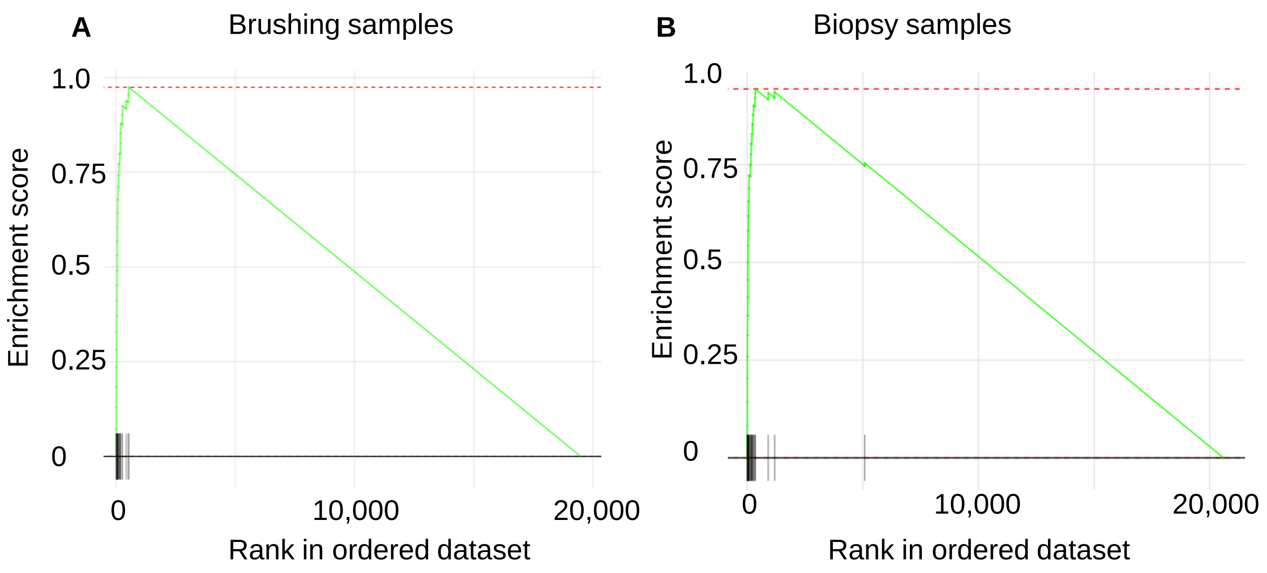


**Supplementary Figure E9**

Geneset enrichment analysis profiles amongst fluticasone treatment-related differentially expressed genes in healthy controls, showing very strong enrichment for a set of 26 genes shown to be induced in participants with asthma by 10 weeks of inhaled fluticasone 500 mcg BD in a microarray analysis of epithelial brushings by Woodruff *et al*(E20). NES, normalised enrichment score. Adjusted P<<0.0001.

### Supplementary Figure E10

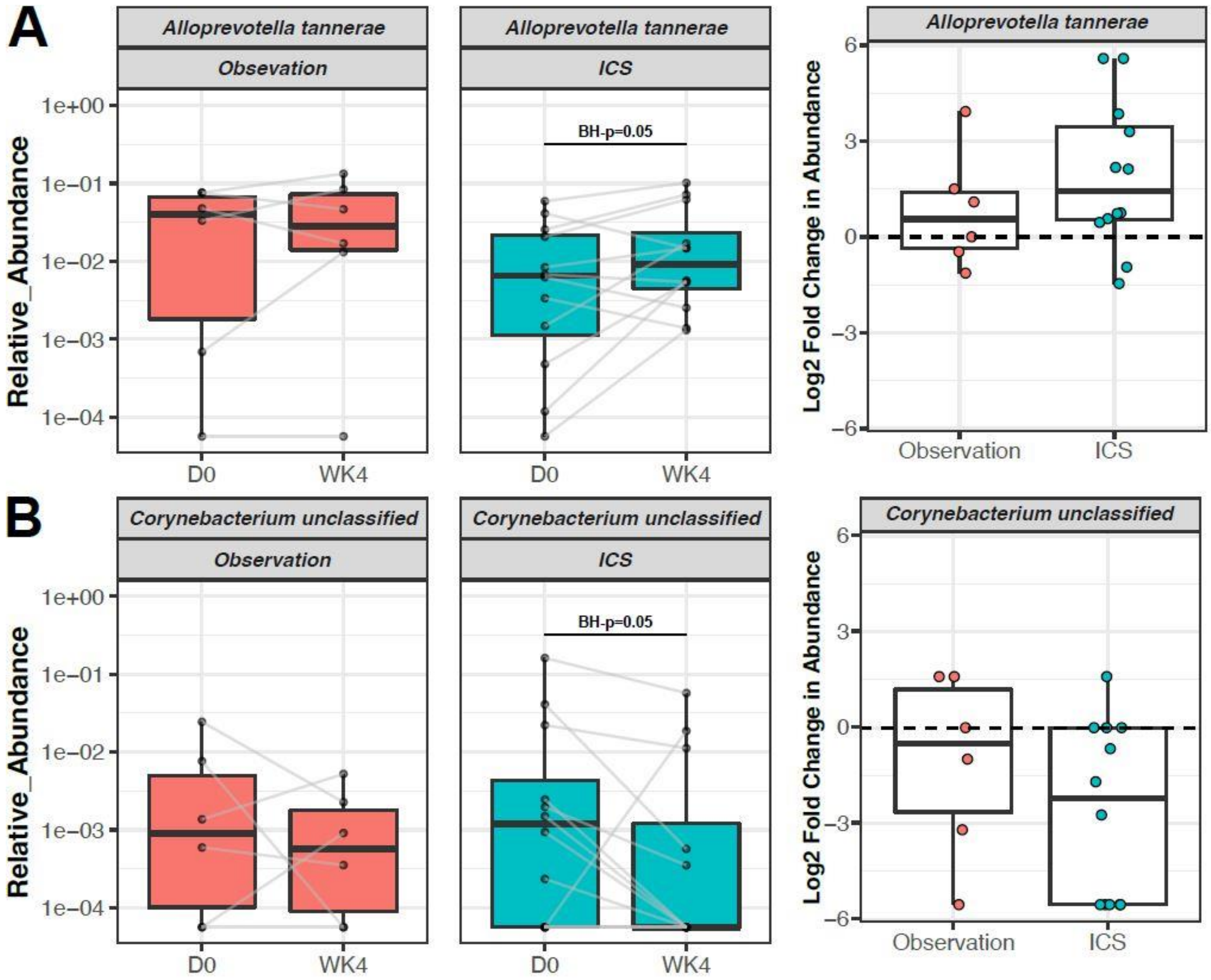


**Supplementary figure E10**. Changes in **A**) Alloprevotella tannerae and **B**) Corynebacterium unclassified after 4 weeks of ICS-treatment or observation, which were significant with Benjamini-Hochberg adjusted p-value < 0.05, expressed as log2 fold change in relative abundance of taxa.

### Supplementary Figure E11

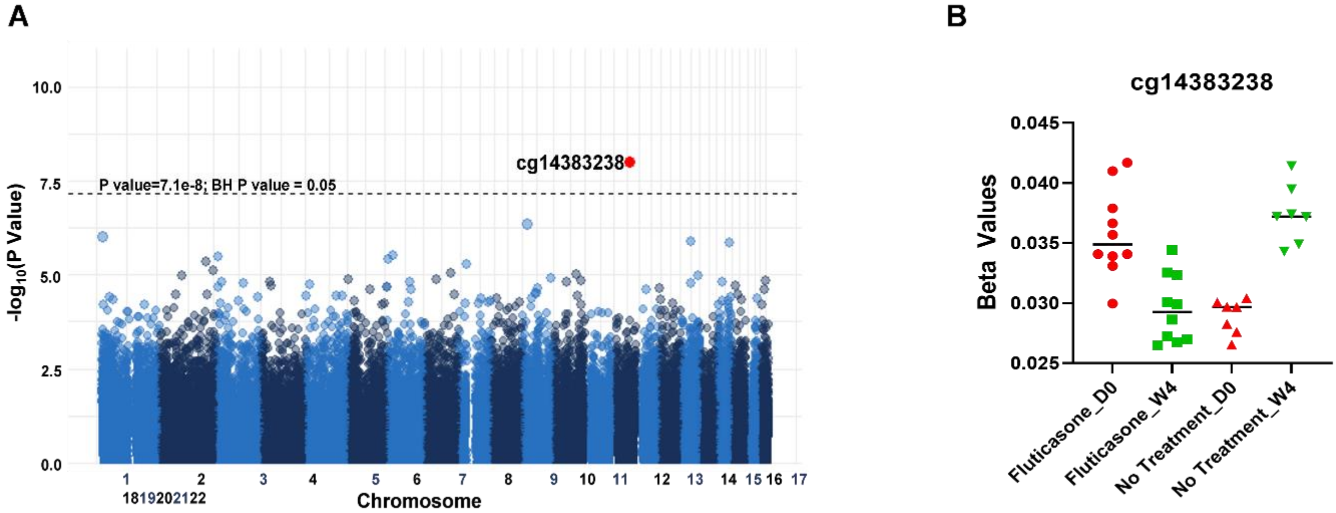


**Supplementary Figure E11. A**) Manhattan plot depicting the results of the linear regression interaction model investigating the effect of fluticasone treatment for 4 weeks. Genomic location of CpG sites are represented on x-axis and -$\log_{10}$(p values) are represented on the y axis. The highlighted point (red) represents the only CpG site (cg14383238) that was significantly differentially methylated at week 4 compared to day 0 in response to fluticasone treatment. **B**) Contrasting trends observed in cg14383238 methylation between week four and day zero of fluticasone-treated and untreated groups. (D0- Day 0; W4- Week 4).

399
